# A randomized controlled trial of postbiotic administration during antibiotic treatment increases microbiome diversity and enriches health associated taxa

**DOI:** 10.1101/2024.07.25.24311015

**Authors:** Jonas Schluter, Fanny Matheis, Wataru Ebina, William Jogia, Alexis P. Sullivan, Kelly Gordon, Elbert Fanega de la Cruz, Mary E Victory-Hays, Mary Joan Heinly, Catherine S. Diefenbach, Jonathan U. Peled, Kevin R. Foster, Aubrey Levitt, Eric McLaughlin

## Abstract

Antibiotic-induced microbiome injury, defined as a reduction of ecological diversity and obligate anaerobe taxa, is associated with negative health outcomes in hospitalized patients, and healthy individuals who received antibiotics in the past are at higher risk for autoimmune diseases. No interventions are currently available that effectively target the microbial ecosystem in the gut to prevent this negative collateral damage of antibiotics. Here, we present the results from a single-center, randomized placebo-controlled trial involving 32 patients who received an oral, fermentation-derived postbiotic alongside oral antibiotic therapy for gastrointestinal (GI)- unrelated infections. Postbiotics comprise complex mixtures of metabolites produced by bacteria during fermentation and other processes, which can mediate microbial ecology. Bacterial ecosystem alpha diversity, quantified by the inverse Simpson index, during the end of the antibiotic course was significantly higher (+40%) across the 16 postbiotic-treated patients compared with the 16 patients who received a placebo, and the postbiotic was well-tolerated. Secondary analyses of 157 stool samples collected longitudinally revealed that the increased diversity was driven by enrichment in health-associated microbial genera: obligate anaerobe Firmicutes, in particular taxa belonging to the Lachnospiraceae family, were higher in treated patients; conversely, *Escherichia/Shigella* abundances, which comprise pathobionts and antimicrobial-resistant strains, were reduced in postbiotic-treated patients at the end of their antibiotic course and up to 10 days later. Taken together, these results indicate that postbiotic co-administration during antibiotic therapy could support a health-associated gut microbiome community and may reduce antibiotic-induced microbiome injury.

- Postbiotic administration during antibiotic treatment increases bacterial alpha diversity
- Characteristic bacterial signatures are associated with postbiotic administration
- Health-associated taxa are enriched and disease-associated taxa are reduced by postbiotic treatment

## Introduction / Background

Antibiotic therapy is the most effective treatment to fight bacterial infections and indispensable in modern medicine; it can cure mild disease and save lives in severe cases of bacteremia^1^, organ invasion and sepsis^2^. Recently, however, evidence has accumulated that oral antibiotic treatment may collaterally injure the human microbiome^3–11^. Antibiotics may interfere with commensal bacterial growth, or can kill health-associated bacterial populations ^3,9^, a phenomenon that may have been vastly underestimated^3^. Such disruption of the commensal microbial ecosystem can persist for months following antibiotic exposure^7^ and is seen in otherwise healthy individuals^12^ as well as severely immunocompromised, hospitalized patients who, in some instances, lose most of the normally resident bacteria^8–11^. Clearance of the normal flora can enable persistent colonization by pathogens, e.g., *Clostridioides difficile*, and cause recurrent, difficult-to-cure, infections^13,14^.

Antibiotic-induced interference with bacterial reproduction and killing of gut bacterial populations lead to shifts in the gut microbiome ecosystem that can be characterized by a decline in alpha diversity^9,10,15^. Alpha diversity metrics are summary measures quantifying the ecological composition of a microbiome sample; they may capture the number of taxonomic groups (e.g. more taxa increase diversity) and their abundance distributions (e.g. more evenly distributed taxon abundances increase diversity)^16^. Therefore, they have been used to quantify microbiome injury at the ecosystem level^17^. Notably, antibiotic use and loss of bacterial microbiome alpha diversity have been associated with health risks, including higher mortality in hematopoietic stem cell transplantation^18,19^, translocation of microorganisms from the gut into the bloodstream (often resulting in bacteremia and sepsis) in immunocompromised patient populations^10,11,15,20,21^, increased risk for inflammatory bowel disease^22^ and severity of autoimmune and allergic diseases^23–27^. Furthermore, loss of bacterial diversity can affect cross-kingdom interactions: depletion of bacterial competitors may allow for fungal species to expand^28^, and fungal expansions were associated with increased risk of fungal bloodstream infections in cancer patients^15^ as well as fungal urogenital infections in otherwise healthy women^29^. Recently, antibiotic exposure has been associated with worse response to cancer immunotherapies^30–34^, and direct links between perturbation of the microbiome and human immune system modulation are emerging^35–37^.

A diverse microbiome, predominantly populated by obligate anaerobe taxa, is thus associated with good health. As a result, means to restore the microbiome following antibiotic exposure are much sought after. Autologous fecal microbiota transplantation (auto-FMT) can recover antibiotic-depleted bacteria using fecal materials harvested and stored prior to treatment^38^. Heterologous FMTs with material from healthy donors (allo-FMT) have shown promising results in improving inflammatory bowel disease^39^, response to immune therapy^40^, and successfully reduced *C. difficile* abundances and recurrence^41,42^. FMTs, however, come with risks^43^: antimicrobial-resistant strains are prevalent in the microbiome^11^ and can sometimes cause life-threatening complications during FMT^44^. Furthermore, matching donor to recipients for optimal engraftment remains an unsolved challenge^45^. Nevertheless, recently, the U.S. Food and Drug Administration has approved two FMT-based therapies: a rectally administered single dose of prepared stool from healthy donors and a fecal microbiota-based oral treatment, both to prevent the recurrence of *C. difficile* infection^46,47^. In addition to large community transfers via FMT, administration of selected live microbial species, termed probiotics, have been established, albeit with conflicting data on efficacy^48,49^, and variable recipient colonization^50,51^. Whilst probiotics are generally assumed to be safe, their use has been associated with adverse outcomes^48^, such as bacteremia^52^, fungemia^53^, and bowel ischemia^49^ especially in immunocompromised and critically ill patients. Facilitation of probiotic colonization following antibiotic-induced depletion of recipient commensals can also lead to persistent dysbiosis and delay recovery of the pretreatment microbiome^54^. These shortcomings of probiotics highlight the potential benefit of modulating resident commensal microbial ecology without the addition of new microbial strains and species towards safe and robust microbe-targeted therapies. Aside from live therapeutics, one potential alternative is the use of prebiotics. These are substances intended to provide nutrients to specific, desired microbial populations, and have recently been shown to have some effects on microbiome communities^37^. Surprisingly, however, a beneficial, anti-inflammatory effect was observed resulting from increased consumption of fermented foods^37^. Thus, products of fermentation may offer a novel therapeutic avenue to manipulate the microbiome.

As such, an emerging alternative to live therapeutics, which we explore here, is the use of postbiotics. Postbiotics is an umbrella term that comprises complex mixtures of metabolites produced by bacteria for example during fermentation, as well as cell wall components and other dead cell components (as well as some potentially viable cells^55^).

Emerging pre-clinical and clinical studies indicate health-beneficial effects of postbiotics^37,56,57^. Thus, postbiotics may offer a safe novel therapeutic avenue to target the microbiome. In particular, we hypothesize that reduction of antibiotic-induced microbiome damage could prevent antibiotic-induced impairment of novel immunotherapies and health sequalae^30,58^. However, it is not yet known if postbiotics can reduce microbiome diversity loss during antibiotic treatment. Here, we describe the results of a single-center randomized controlled trial to assess the ability of a fermentation-derived postbiotic to prevent antibiotic-induced injury to the commensal microbiome in patients receiving oral antibiotics. Following a course of antibiotics, we collected stool samples at five timepoints and profiled the composition of bacterial populations. Our data indicate that treatment with a postbiotic during a course of oral antibiotics supports gut microbial ecosystem stability and health-associated taxa.

## Results

### Overview of study design and intervention

A single-center, randomized placebo-controlled trial of postbiotic treatment (T, n=16) versus placebo (C, n=16) was conducted to measure bacterial alpha diversity as a primary endpoint at the end and directly after finishing a course of oral antibiotics. The goal was to determine feasibility, safety, and a preliminary efficacy assessment of the procedure for prevention of medication-induced injury of the gut microbiota (Study title: Randomized controlled trial of PBP-GP-22 to affect microbiome composition; https://www.isrctn.com; trial registration ISRCTN30327931). Otherwise healthy ambulatory adult patients (**Table 1**) presenting to a single clinical site in Houston, Texas, between 05 and 06/2018 for non-gastrointestinal infections were recruited. After giving informed consent, patients were provided with take-home stool sample collection kits, a supply of a commercially available probiotic, and a supply of the placebo (control, C) or postbiotic (treatment, T) to be taken during and ten days beyond the antibiotic course (administered antibiotics are listed in **Table S1**) along with the commercial probiotic and their standard-therapy antibiotic (**Figure 1, S1**, CONSORT report^59^). We chose to include the probiotic background, which included six strains of *Lactobacillus* and *Bifidobacterium*, because it was the standard recommendation by the participating physicians when administering antibiotics. Moreover, by including an existing treatment in our study, our design represented a relatively stringent test of the ability of a postbiotic to further improve outcomes. A total of 157 stool samples were collected longitudinally, for up to five timepoints (S1-S5) relative to the end of a full course of prescribed antibiotics per patient (**Figure 1A**).

**Figure 1:**
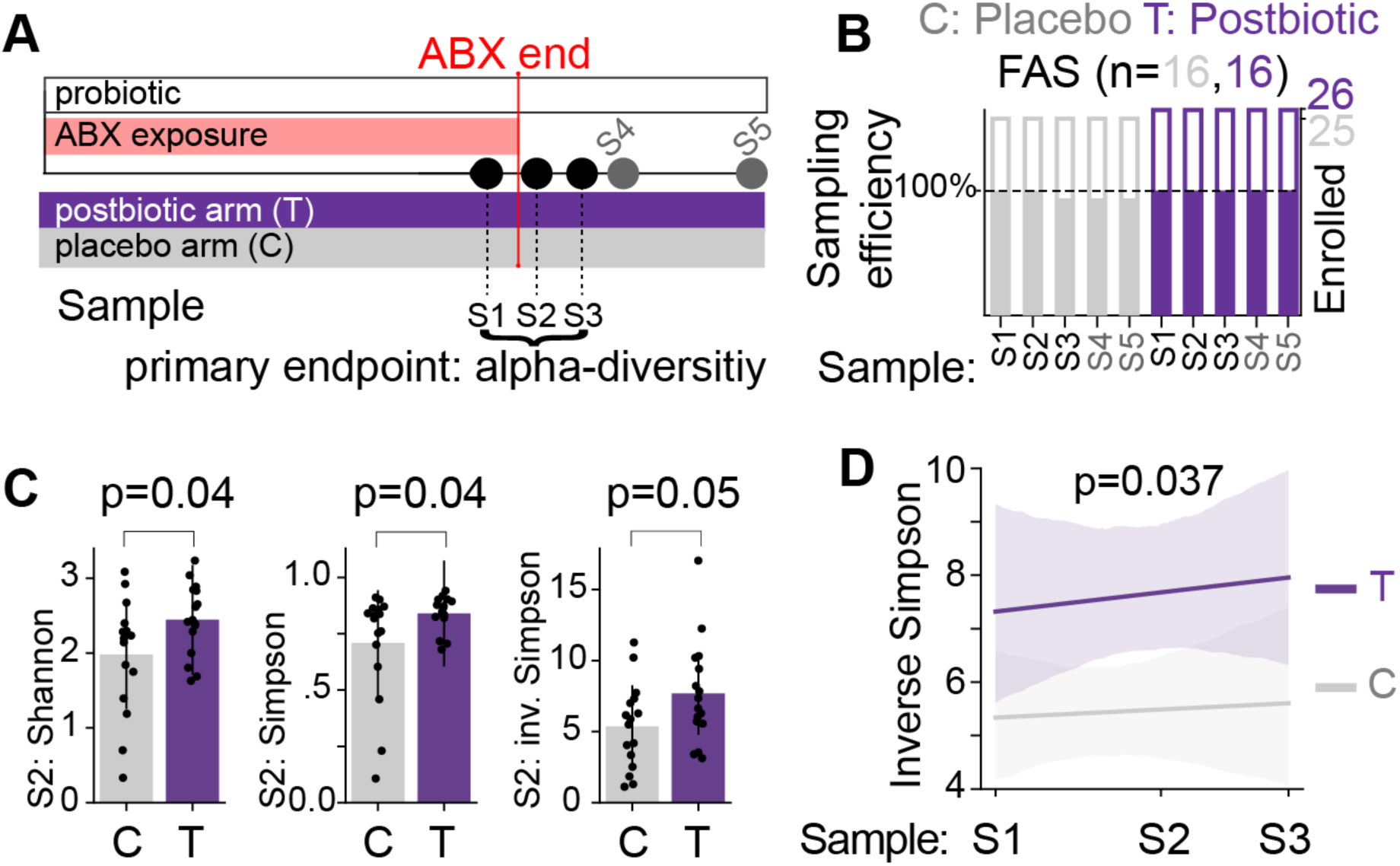
Postbiotics significantly increased bacterial alpha diversity at the end of an antibiotic course. **A)** Sample collection protocol and study design; primary endpoint of the trial was alpha-diversity (inverse Simpson index) during the end of the antibiotic course (S1-S3), additional endpoints were alpha-diversity (inverse Simpson) at later time points (S4, S5). **B)** Enrolment and sample collection efficiency in percent of planned vs obtained; Full Analysis Set, (FAS): 100%=16 samples per arm; cohort report in **Figure S1**. **C)** Alpha diversity directly after finishing the antibiotic course (S2, p-values from two-sided t-tests). **D)** Inverse Simpson alpha diversity across timepoints S1-S3 was significantly higher in treated (purple) than control (grey) patients, see also **Figure S2**; p-value from linear mixed effects model^38^.

**Table 1:** Patient characteristics and indications by treatment arm, with statistical tests for *p* value calculations indicated.

| Characteristic |  | Control | Treatment | *p* value |
| --- | --- | --- | --- | --- |
| Age |  | 39, [20,60] | 45, [20, 64] | 0.95 (t-test) |
| Sex | Female | 10 | 8 | 0.72 (Chi2) |
|  | Male | 6 | 8 |  |
| Disease |  |  |  |  |
|  | Sinusitis | 5 | 3 |  |
|  | Cellulitis | 2 | 1 |  |
|  | Kidney infection/UTI | 1 | 0 |  |
|  | Abscess | 1 | 0 |  |
|  | Ear infection | 2 | 0 |  |
|  | Cyst | 1 | 0 |  |
|  | Pharyngitis/Strep | 1 | 6 |  |
|  | Bronchitis | 1 | 2 |  |
|  | Laceration | 0 | 1 |  |
|  | Toothache | 0 | 1 |  |
|  | Other | 2 | 2 |  |

It was hypothesized that microbiome injury would be maximal at the end of a completed course of antibiotics. Therefore, the primary endpoint was set as bacterial alpha diversity, measured by the inverse Simpson index on three timepoints at the end of the antibiotic course: on the day prior to finishing the antibiotic treatment (S1), directly after completing the antibiotic (S2) and one day thereafter (S3). Among patients who entered the study, stool sampling rates were high, with only a single sample from S3 missing in the final analysis set (FAS, target participant number: 25, FAS: 32, **Figure 1B**). Patients were given a phone number to call to report any side effects, and were contacted via text messages prior to fecal sample collection; there were no grade 3 or higher side effects reported (**Table S2**).

### Bacterial alpha diversity at the end of an antibiotic course is high in patients receiving the postbiotic

Microbiome profiling by amplification and sequencing of the V4 region of the 16S rRNA gene was performed on DNA collected from 157 samples. Bacterial alpha diversity measured at the end of the antibiotic treatment (S2) was significantly higher in postbiotic-treated patients compared with placebo recipients for different alpha diversity metrics (**Figure 1C**, Shannon: +24%, p=0.046; Simpson: +19%, p=0.048; inverse Simpson: +43%, p=0.056, t-test). Time series analysis^38^ of longitudinally collected samples (**Figure S2**) showed that postbiotic-treated patients had significantly higher alpha diversity during the end phase of their antibiotic treatment (**Figure 1D**, p=0.037, linear mixed effects model). Together, these results indicate that addition of this postbiotic to an orally-administered antibiotic course is clinically feasible, well-tolerated, and significantly increased gut microbial alpha-diversity.

### Postbiotic treatment is associated with characteristic bacterial signatures

We next sought to analyze differences between microbial ecosystems in treated vs control patients at a higher resolution than resolved by alpha diversity metrics. For this, we first visualized the individual patients’ compositional time courses by plotting the relative abundances of the most abundant bacterial families (**Figure 2A**), which indicated compositional variation across patients and changes over time in several individuals. To analyze compositional variability, we next performed a principal coordinate analysis (PCoA) on sample-by-sample Bray-Curtis distances calculated on operational taxonomic units’ (OTUs) relative abundances (**Figure 2B-E**). Samples from patients of either sex localized across the projected bacterial compositions (**Figure 2B**). In most samples, obligate anaerobe Bacteroidetes taxa represented the most abundant bacterial families (**Figure 2C**), whereas samples from both experimental arms contained only low relative abundances of the genera corresponding to the probiotics taken by all patients (median relative genus abundance: *Bifidobacterium* [C: 3.9e-4, T:4.3e-4], *Lactobacillus* [C: 1e-4, T: 0.9e-4]; **Figure S3**). Consecutive samples from the same patient tended to localize near one another, with significantly lower intra- than inter-patient variability during the transition from antibiotics into the post-antibiotic period (**Figure 2D**), even though the observed microbiome dynamics in several patients spanned wide regions of the compositional space.

**Figure 2:**
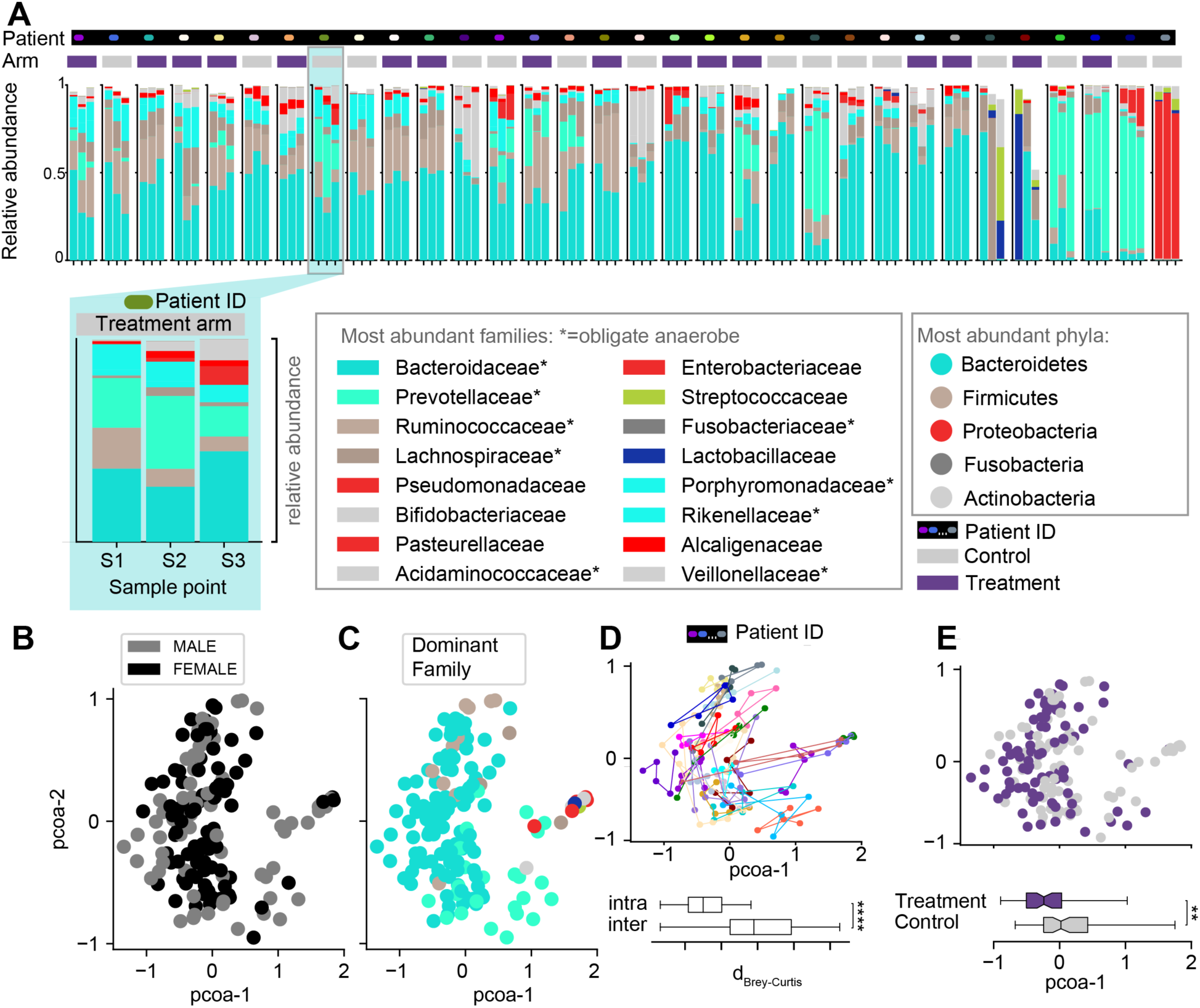
Bacterial compositions in longitudinally collected fecal samples. **A)** Bacterial family compositions in samples S1-S3 for each patient; families of identical higher taxonomic groups in similar colors, one patient time course enlarged. Patients are sorted by average alpha diversity across samples. Patient ID (colored dot on black background) and treatment arm (grey box: control, purple box: treatment) are indicated above the time courses. **B-E)** PCoA plots of bacterial relative abundances in all 157 samples, colored by sex (**B**), most abundant bacterial family per sample (**C**, colors as in A). **D**) Patients’ compositional trajectories visualized on the PCoA plot (colors as in A), with boxplot showing intra- and inter-subject Brey-Curtis dissimilarities. **E)** PCoA labeled by treatment, with significant differences between the groups along the first principal coordinate indicated (**: p<0.01, ****:p<10-4, two-sided t-test).

While samples from treated and control patients localized across the PCoA space, a statistical analysis indicated that the samples from treated patients varied consistently in their composition from those observed in control patient samples (**Figure 2E**), which suggested characteristic gut bacterial communities in treated patients.

### Postbiotic treatment is associated with an enrichment in health- and a reduction in disease-associated taxa

To analyze which taxa were consistently higher in treated patients or lower alongside the reduced diversity in control patients, we compared the relative abundance profiles between the two patient groups directly after finishing the antibiotic course (S2), and over time (S1-S3). To account for the compositional nature of 16S relative abundance data, we calculated the centered-log ratio (CLR) transformation of relative taxon abundances across all samples. To reduce sparsity of the OTU-level data, we aggregated relative abundances at the phylum, family or genus levels prior to CLR transformation.

Statistical comparison of the CLR-transformed phylum relative abundances directly after finishing the antibiotic course (S2) revealed a significantly higher abundance of Firmicutes among treated patients (**Figure 3A**, p-value=0.002, q-value<0.05, 0.1 FDR). At higher taxonomic resolutions, we analyzed the 10 most abundant bacterial families and 20 most abundant genera after CLR-transformation: Lachnospiraceae and Bacteroidaceae (family-level analysis), and *Roseburia*, *Bacteroides*, and *Ruminiclostridium* 5 (genus-level analysis) were enriched in treated patients at S2 (**Figure 3B, C**). However, the observed taxon enrichments, while significant in univariate analyses, did not remain significant after correcting for multiple hypotheses, with the exception of the observed enrichment of Lachnospiraceae in treated patients (univariate p-value=0.004, q<0.05, 0.1 FDR), which remained significant.

**Figure 3:**
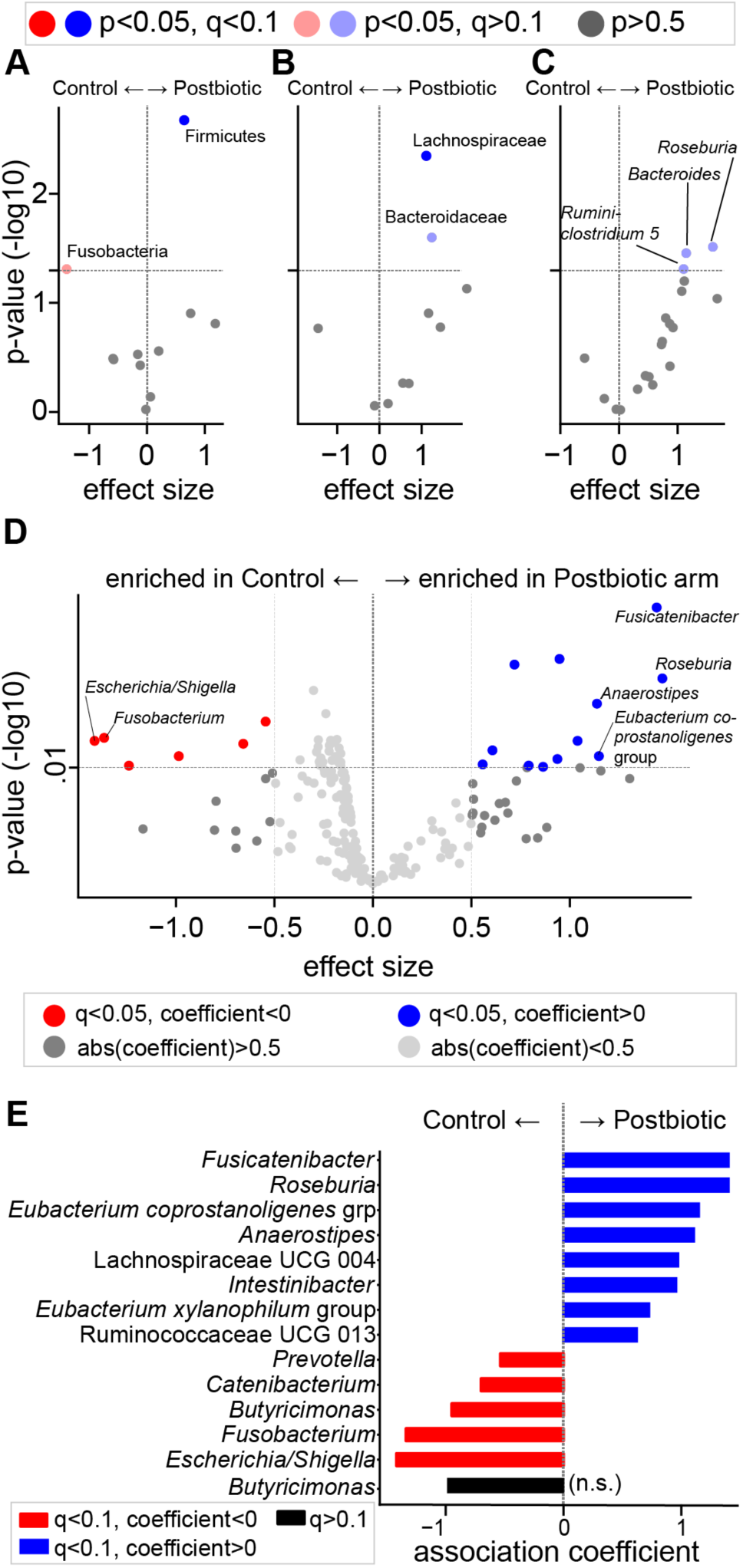
Health-associated taxa are enriched in postbiotics treated patients. V**A-C)** olcano plots of association coefficient effect sizes between CLR-transformed taxon abundances and treatment arm; control (left), treatment (right) in samples collected at the end of an antibiotic course (S2). Negative log_10_- transformed p-values of association coefficients from 10 most abundant phyla (**A**), families (**B**), and 20 most abundant genera (**C**); significant associations in color (light color: univariate p-value<0.05, full color: q-value<0.05, 0.1 FDR, Benjamin/Hochberg method). **D)** Volcano plot of associations between CLR-transformed relative genus abundances with treatment or control in samples S1-S3 (100 most abundant genera analyzed). **E**) Significant genera from D re-analyzed with a mixed effects model using random intercepts per patient to account for multiple samples per patient; colored bars indicate significant association coefficients (red: higher in control, blue: higher in treatment arm) after multiple hypothesis correction via the Benjamin/Hochberg method (FDR: 0.1, q<0.05).

We next analyzed CLR-transformed genus abundances in samples S1 to S3. Using a more stringent false discovery rate (0.05 FDR) as well as a minimal effect size cutoff (0.5), revealed several genera significantly enriched in treated patients, including *Anaerostipes*, *Eubacterium coprostanoligenes* group, *Fusicantenibacter*, and *Roseburia*, and significantly reduced genera in treated patients, including *Fusobacterium* and *Escherichia/Shigella* (**Figure 3D**, **S4**). We tested the 13 differentially abundant genera in a mixed effects model (**Figure 3E**), which confirmed the associations found in the univariate pooled analysis screen (**Figure 3D**). Furthermore, consistent with these results, we found similar associations when performing multivariate analyses using regularized regression approaches (**Figure S5**), which confirmed a trend towards enrichment of commensal obligate anaerobe genera among treated patients. We also compared the abundances of anaerobe taxa discussed as immunomodulatory in the context of cancer therapies^31,32,35^, and found higher abundances of Lachnospiraceae, which comprise *Faecalibacterium*, Ruminococcaceae, which comprise *Ruminococcus*, and Verrucomicrobiaceae, which comprise *Akkermansia*, in treated patients (**Figure S6**). Interestingly, this enrichment of health-associated anaerobe taxa and concurrent lower levels of facultative anaerobe, pathobiont- comprising taxa in treated patients appeared to persist beyond the immediate end of the antibiotic exposure (**Figure S7**), even though alpha diversity in control patients had already approached similar levels to treated patients ten days after the antibiotic course ended (**Figure S8**). In particular, facultative-anaerobe gamma-proteobacteria of the *Escherichia/Shigella* genus were reduced in treated patients even 10 days after finishing their antibiotic course (**Figure S7B**).

Taken together, these results support an enrichment of a health-associated microbial community in patients treated with a postbiotic during their antibiotic course.

## Discussion

We here presented results from a single center randomized placebo-controlled trial to assess the protective effect of a postbiotic adjuvant on microbiome diversity during antibiotic therapy.

Collateral injury to the resident gut microbiota caused by antibiotics has been described as a loss of bacterial ecosystem diversity^18^, loss of commensal microbial taxa^9,15^, and increase of pathobionts, which can comprise antibiotic-resistant species^11^. The enrolled patients were ambulatory and treated with oral antibiotics for non-gastrointestinal infections. Thus, the patient cohort studied here were at a lower risk for microbiome injury-induced complications than for example blood cancer patients^11,15,18^. Among those higher-risk patients, overall survival is statistically associated with antibiotic induced loss of diversity^18^, and response to chimeric antigen receptor T cell therapy is negatively associated with antibiotic exposure^30,34,58^. In hospitalized COVID-19 patients, another high-risk patient group, a combination of antibiotics and viral infection were found to predispose patients to gut born bacteremia^10^. Nevertheless, even in otherwise healthy individuals such as were studied here, post-antibiotic complications such as diarrhoea^60^, post-antibiotic *Clostridioides difficile* infection^61^, or vaginal candidiasis^62^ are common, and selection for antimicrobial resistant strains (AMR) in the gut microbiome may pose a potential health risk during future therapies. Therefore, reducing the negative collateral impact of antibiotics on the microbiome constitutes a critical unmet need.

Our results indicate that microbiome injury is lower when co-administering an oral postbiotic alongside the oral antibiotic: bacterial alpha diversity was significantly higher at the end of an antibiotic course in patients who received the postbiotic instead of the placebo, and this increased diversity was driven by commensal taxa. Health-associated obligate anaerobe organisms were higher in treated patients: *Fusicantenibacter*, which is depleted in rheumatoid arthritis patients^63^, short-chain fatty acid-producing *Roseburia*, one of the most abundant genera in the healthy gut microbiome^64^, the butyrogenic *Anaerostipes* genus, which comprises lactate- consuming species^65^, and the *Eubacterium coprostanoligenes* group, which has been hypothesized to be involved in beneficial fat metabolism by reducing cholesterol^66^. Conversely, *Escherichia/Shigella*, a genus of gram negative facultative anaerobe species that comprise multiple antibiotic-resistant strains and pathobionts which can cause secondary bacteremia when translocating across a weakened gut barrier^10,11^, were significantly lower in treated than in control patients. Interesting, we also observed that *Fusobacterium*, an obligate anaerobe taxon, was significantly lower in treated patients. Its predominant species, *F. nucleatum*, is commonly found in the oral microbiome and associated with the development of colorectal cancer when abundant in the gut^67,68^. It is plausible that the higher diversity of commensal gut bacteria in treated patients improved colonization resistance and reduced gut colonization of *F. nucleatum* when swallowed. Furthermore, the higher diversity of commensal organisms may have suppressed the opportunistic expansion of *Escherichia/Shigella* observed in control patients via competitive exclusion^11,69–71^.

Limitations of the study include that both treatment arms received a commercial probiotic; however, low abundances of the taxa from the probiotics in both treatment arms indicated that this had no influence on the diversity differences observed between treatment arms. Nevertheless, future studies are warranted to validate the efficacy of the oral postbiotic outside of the context of concurrent probiotic administration. Furthermore, due to the enrollment at an urgent care clinic where patients were instructed to start their treatment immediately, no baseline stool sample prior to antibiotics was collected. However, we speculate that the impact of antibiotics on the microbiome are maximal at the end of an antibiotic course, and that thus the increase in diversity observed among treated patients could reflect protection of the commensal microbiome.

This is a plausible scenario because of the enrichment of commensal taxa in the microbiome of patients treated with the postbiotic, which did not include these taxa. The study results, while significant and of considerable magnitude, are based on a small cohort and the direct impact on patients’ health was not assessed in this trial, which focused on microbiome diversity as a primary outcome. Additional studies are now warranted to validate the herein described beneficial impact of postbiotic treatment on the microbiome and assess if the stabilization of the microbiome translates into a patient health benefit.

Recently, consumption of fermented plant-based foods were described to have a beneficial effect on the microbiome and immune system^37^. The postbiotic under investigation is a complex biologic product derived from the fermentation of medicinal plants by generally regarded as safe (GRAS) bacterial species. Unlike in fermented foods, manufacturing of postbiotics generally includes a dedicated inactivation step that reduces live cell counts^55^, and therefore, they are assumed to have a generally favorable safety profile over live therapeutic based approaches. For the manufacturing of the postbiotic used in this trial, a fermentation process by specific species of *Lactobacillus* and *Bifidobacterium* was used, which represent major taxa commonly present during the assembly of the infant microbiome^72^. While the mechanism of action of the observed protection of gut microbiome diversity is not fully understood, we hypothesize that bacterial metabolites of these species, their products of secondary metabolism and signaling molecules may support or elicit competitive responses within resident members of the commensal adult microbiome, which have evolved to ecologically succeed early colonizers of the gut microbiome^73^, increasing their resilience to external perturbations^74^.

In conclusion, here we have presented evidence that co-administering a fermentation derived postbiotic can increase gut microbiome diversity during antibiotic treatments. The increased diversity was associated with higher abundances of health-associated bacterial taxa, while genera comprising pathobionts were lower among postbiotic treated patients.

## Methods

### Human subjects

**Table 1** reports metadata, including age and sex, corresponding to the 32 healthy adult subjects enrolled in a single site double-blind randomized controlled study (trial registration ID: ISRCTN30327931). This study was reviewed and approved by New England Institutional Review Board, an independent IRB located in Needham, MA. All subjects were provided with an explanation of the study and gave informed consent in writing prior to the start of their participation in the study.

### Human subject study methods

Fifty-one ambulatory adult subjects were initially evaluated for eligibility (see inclusion and exclusion criteria used to evaluate subject suitability listed below), one was not eligible. Fifty of the subjects were then enrolled in a single site double-blind interventional study with a randomized-controlled design to test the hypothesis that patients receiving an oral postbiotic would present with higher bacterial alpha diversity, measured by the inverse Simpson index, during three time points at the end of an antibiotic course than patients receiving a placebo control. Analyzed timepoints were: the final day of antibiotics, the first day after completing the antibiotics course, and the second day after completing the antibiotics course. Analysis was performed by time series analysis that accounts for repeated measurements and uses time as categorical predictors, as done before^38,75^. This study was reviewed and approved by New England Institutional Review Board, an independent IRB located in Needham, MA. Eligibility for enrollment was determined by the following inclusion and exclusion criteria:

Inclusion criteria:

- Otherwise healthy adults with a body mass index (BMI) (18-28 kg/m²)
- Patients are prescribed a course of antibiotics with a duration of at least 5 days
- Patients have not had another course of antibiotics in the past 6 months
- Patients are willing to cease taking any other supplements Exclusion criteria:
- patients suffering from gut related illness (regular and severe constipation, diarrhea, inflammatory bowel disorder or inflammatory bowel disease).
- patients who usually experience severe diarrhea (e.g. for more than 3 days) following antibiotic treatment.
- patients who have a compromised immune system, are taking any immune modulators or have an autoimmune disorder.
- patients who are diabetic or take any blood pressure medication.
- patients who have any known allergies to the probiotics, or the herbs and bacteria used in the postbiotic formula.
- patients who are pregnant or nursing.

The double-blind randomization scheme for this study was as follows. Patients were given an unmarked box (the study kit) containing stool sample materials, and either control or treatment materials neither of which were marked. To blind study coordinators, boxes had been shuffled after assembly of all study kits, prior to the study. Kits contained barcodes that allowed identification of participants’ treatment arm after the participant handed over their samples. One of the subjects evaluated for participation was ineligible due to not meeting the inclusion criteria. Fifty subjects received the study kits. Seventeen subjects withdrew consent during follow-up; six and twelve subjects from the control and the treatment arm, respectively, without explanation.

One patient from the treatment arm was removed from the study due to a second antibiotic prescription which was not declared during enrolment.

Treatment regimens were as follows. In their study kit, the control arm received a commercially available probiotic in unmarked capsules and a resistant starch placebo in an unmarked capsule; the treatment arm received the same commercially available probiotic in unmarked capsules and the fermentation-derived postbiotic in unmarked capsules. Subjects were instructed to take one capsule of each every day during their antibiotic course and for ten days after finishing the antibiotic course.

Patients were given a phone number to call to report any side effects; no such phone calls were received. Prior to the collection of each stool sample, study coordinators were checking in with patients via text message to remind them of the sample collection and ask for side effects, collectively presented in **Table 2**. Patients were contacted on study days 1,2,3,4 and 10, and there were no grade 3 or higher side effects reported.

### Eligibility criteria, ethical approval, and consent to participate

This study protocol was approved by the institutional review board (IRB) number: 120180088, trial registration ID: ISRCTN30327931. Otherwise healthy ambulatory adult patients (**Table 1**) presenting to a clinic in Houston, Texas, between 05 and 06/2018 for non-gastrointestinal infections were recruited. Participants were recruited by physicians. Onsite nurses, trained in human subject research, reviewed the consent form with the potential participants and answered any questions about the study and its potential risks. Informed consent was obtained from all participants. Participants were compensated with a $15 Amazon gift card per sample collected at the end of the study. If a participant did not complete the study, they were not penalized and rewarded for the samples they collected.

### Sample collection

After informed consent was obtained, participants were provided with 5 Diversigen OmniGene Gut kits for stool collection at home, a kit which stabilized samples at room temperature for up to 60 days. Patient were handed a work sheet listing their expected samples, recorded the date of sample collection, and shipped all samples in one batch. Samples were received and processed by a specialized contract research organization, Diversigen, (Houston, Texas).

### Postbiotic composition

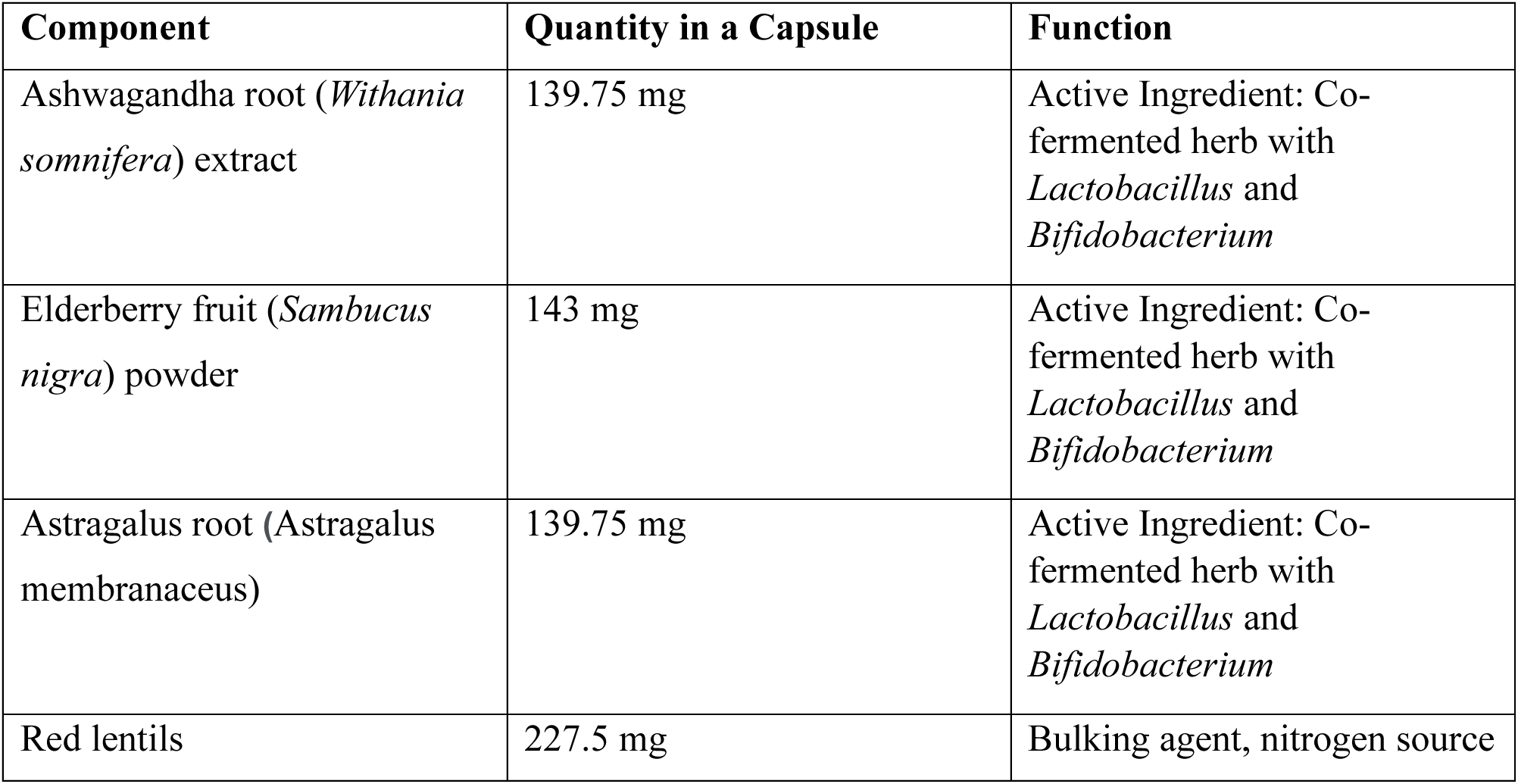

| Component | Quantity in a Capsule | Function |
| --- | --- | --- |
| Ashwagandha root (*Withania somnifera*) extract | 139.75 mg | Active Ingredient: Co-fermented herb with *Lactobacillus* and *Bifidobacterium* |
| Elderberry fruit (*Sambucus nigra*) powder | 143 mg | Active Ingredient: Co-fermented herb with *Lactobacillus* and *Bifidobacterium* |
| Astragalus root (*Astragalus membranaceus*) | 139.75 mg | Active Ingredient: Co-fermented herb with *Lactobacillus* and *Bifidobacterium* |
| Red lentils | 227.5 mg | Bulking agent, nitrogen source |

### DNA extraction and bacterial 16S rRNA gene sequencing

DNA from human samples was extracted with PowerSoil Pro (Qiagen) on the QiaCube HT (Qiagen), using Powerbead Pro (Qiagen) plates with 0.5mm and 0.1mm ceramic beads. All PCR products were analyzed with the Agilent TapeStation for quality control and then pooled equimolar and sequenced directly in the Illumina MiSeq platform using the 2x250 bp protocol. Human samples were prepared with a protocol derived from 44, using KAPA HiFi Polymerase to amplify the V4 region of the 16S rRNA gene. Libraries were sequenced on an Illumina MiSeq using paired-end 2x250 reads and the MiSeq Reagent Kitv2. Sample processing and sequencing were performed by Diversigen.

### Bioinformatic processing and taxonomic assignment

OTU tables were generated by Diversigen bioinformatics pipelines (detailed bioinformatic tools and settings in **Supplementary Methods**). Briefly, Useach v7.0 was used to process raw sequencing reads, and taxonomy was assigned via the Silva database v128.

### Statistical analyses

Unless otherwise stated, statistical analyses were performed with Python 3, version 3.10.4 and R, version 4.2.

We calculated the inverse Simpson (IVS) index from relative ASV abundances (p) with N ASVs in a given sample, 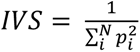. Regression analyses were conducted using the statsmodels package for the Python programming language, using the functions *ols* and *mixedlm* for ordinary least squares and mixed effects modeling, respectively. Relevant code of primary analyses and corresponding data tables are available in the supplementary materials.

To account for compositionality of the relative abundance data, we performed a centered log-ratio (CLR) transformations at different taxonomic aggregation levels, i.e. separate CLR for genus, family, and phylum abundances. While this does not remove compositionality related data analysis challenges, CLR transformations are increasingly used to reduce negative covariance biases in microbiome data. Regression models were implemented using the statsmodels (using functions *ols* and *mixedlm*)^76^ and penalized multiple variate regression models using the sklearn packages (using the function *LogisticRegression*)^77^ for the Python programming language, and using the *nlme*^78^ and *vegan*^79^ library in R.

*p* values were adjusted as appropriate. A *p*-value <0.05 was considered significant after adjustment for multiple comparisons where appropriate. Unless otherwise stated, significance values are noted as follows: * p < 0.05, ** p < 0.01, *** p < 0.001.

Figure 1: Significance of differences in IVS were calculated using a mixed effects model for repeated measures with time treated as a categorical variable^38,75^; significance between arms at individual timepoints was determined using two-sided t-tests.

Figure 2: Differences in sample-by-sample distances were calculated with the adonis2 function from the vegan library for the R programming language.

Figure 3: Single time points were analyzed with univariate linear models using the *ols* function of the statsmodels package, multiple hypothesis correction was performed with the Benjamin/Hochberg procedure using the fdrcorrection function of the statsmodels.stats.multitest library for the Python programming language. The multivariate analysis of significant hits from the univariate screen was performed with the LogisticRegressionCV class from the scikit-learn package for the Python programming language.

## Data availability

Data, statistical test results, and relevant code to reproduce main analyses are available as supplementary materials. The 16S rRNA gene sequencing results are available as supplementary data files (**Table S3**). Raw sequencing reads are available on the sequencing read archive, submission SUB13922145.

## Supporting information

SuppFig1

SuppFig2

SuppFig3

SuppFig4

SuppFig5

SuppFig6

SuppFig7

SuppFig8

SuppMethods

SuppTab1

SuppTab2

## Data Availability

Data, statistical test results, and relevant code to reproduce main analyses are available as supplementary materials. The 16S rRNA gene sequencing results are available as supplementary data files (Table S3). Raw sequencing reads are available on the sequencing read archive, submission SUB13922145.

## Author contributions

AL and EM conceived of the trial, with support by JS. AL, EM, oversaw and conducted the trial, with support by KG, EFC, MEVH, and MJH. JS conducted the analyses and wrote the manuscript with support by CSD, WE, FM, APS, and JUP. JS, FM, WE, WJ, KRF and JUP analyzed and interpreted the results. All authors read and commented on the manuscript.

## Acknowledgements

JS reports funding from the National Institutes of Health (NIH) grants DP2 AI164318-01, and R01CA269617. FM is supported by a Helen Hay Whitney Fellowship. JUP reports funding from NHLBI NIH Award K08HL143189, the MSKCC Cancer Center Core Grant NCI P30 CA008748. We would like to thank Kate Markey for comments and discussion of the manuscript.

## Conflict of Interest Disclosure

AL is CEO of Postbiotics Plus Research. AL and JS are co-founders of Postbiotics Plus Research. AL, JS, EL, KRF, JUP hold equity in Postbiotics Plus Research. JUP reports research funding, intellectual property fees, and travel reimbursement from Seres Therapeutics, and consulting fees from DaVolterra, CSL Behring, and from MaaT Pharma. JUP has filed intellectual property applications related to the microbiome (reference numbers #62/843,849, #62/977,908, and #15/756,845). Memorial Sloan Kettering Cancer Center (MSK) has financial interests relative to Seres Therapeutics. AL and JS have filed intellectual property applications related to the microbiome (reference numbers #63/299,607). JUP and KRF serve on an Advisory board of Postbiotics Plus Research. JS serves on an Advisory board and holds equity of Jona Health.

## Supplementary Material

**Supplementary Figures**

**Figure S1:**
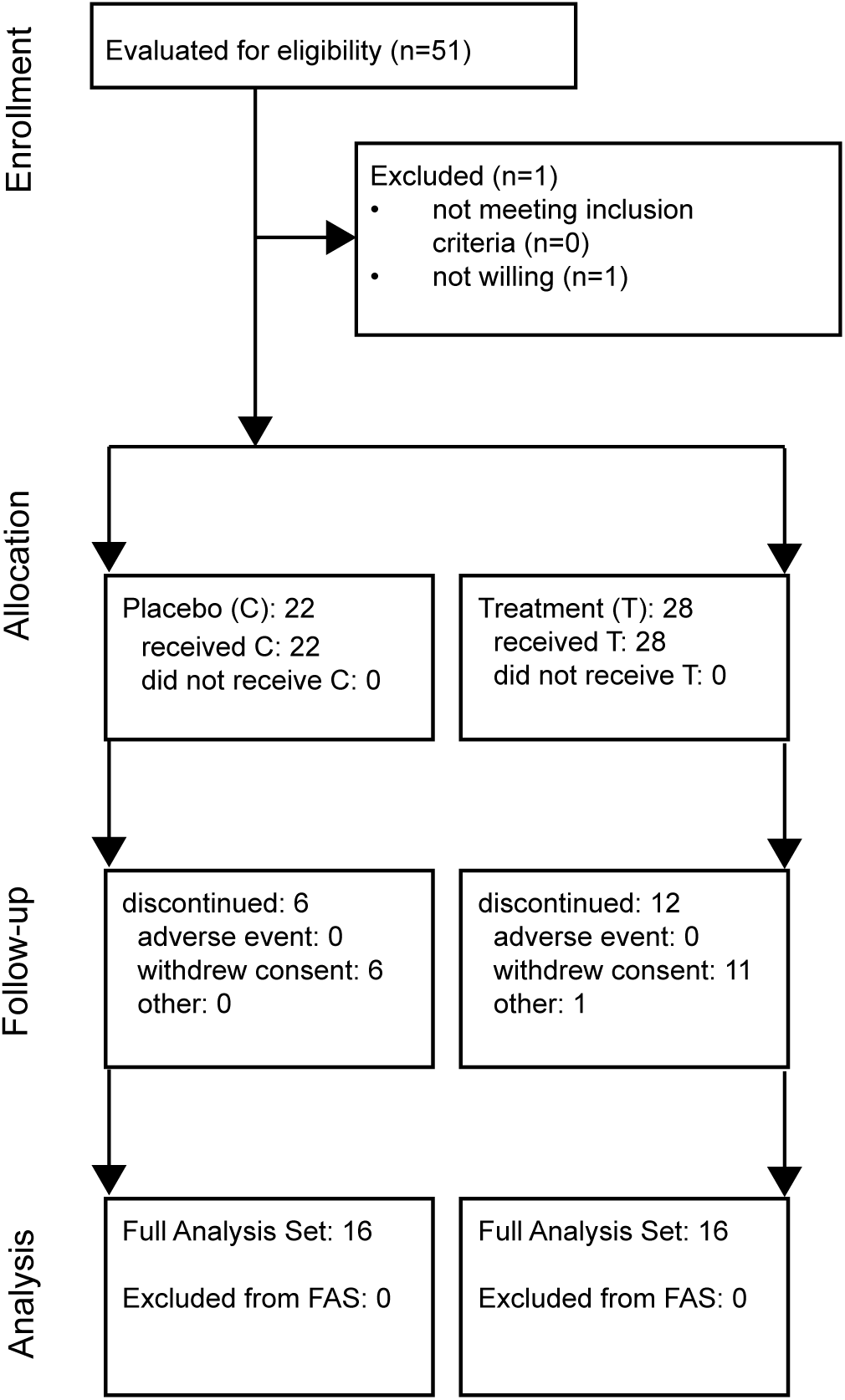
CONSORT Report chart.

**Figure S2:**
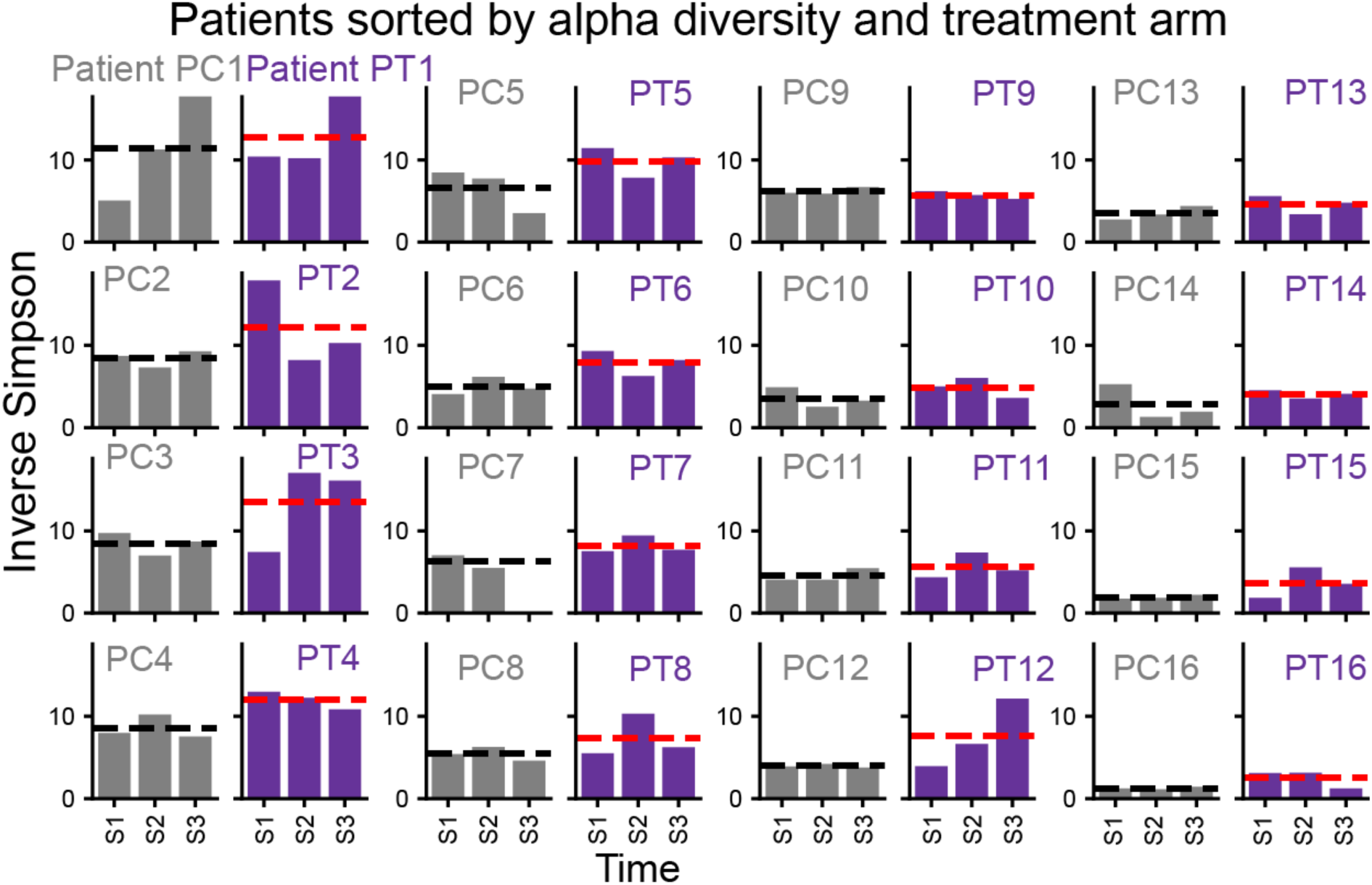
Higher alpha diversity observed across individual patients during primary endpoints (S1-S3). Inverse Simpson alpha diversity in 32 patients (Control: PC, Treated: PT) at the end of an antibiotic course across samples S1, S2, S3; patients sorted by trial arm and average diversity in descending order from top left to bottom right. Dashed lines indicate control (grey) and treated (red) patient averages.

**Figure S3:**
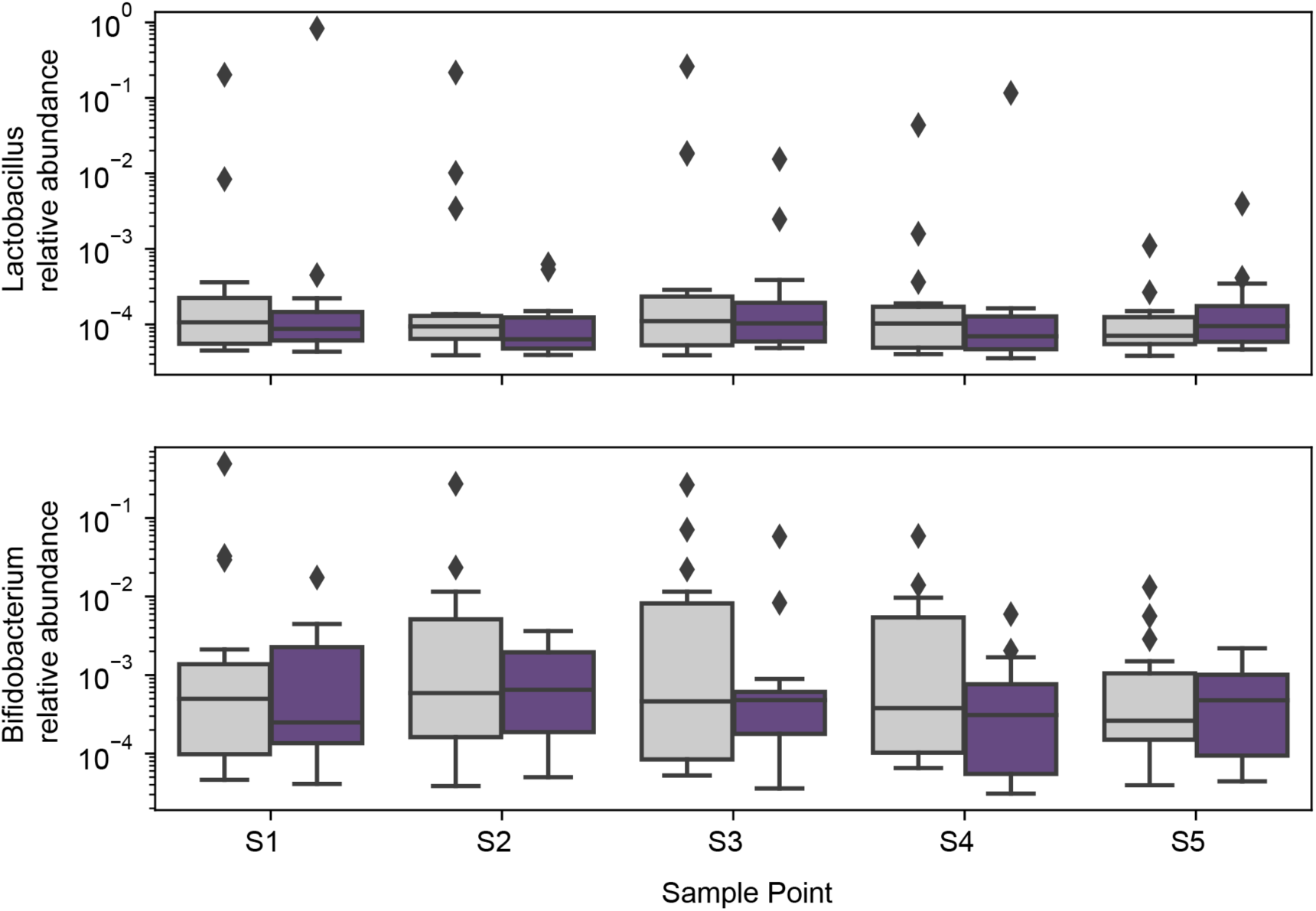
Relative abundances of probiotic genera. Relative genus abundances shown for samples from control (grey) and treated (purple) patients; differences were non-significant at each time point between treatment arms (Wilcoxon rank sum tests).

**Figure S4:**
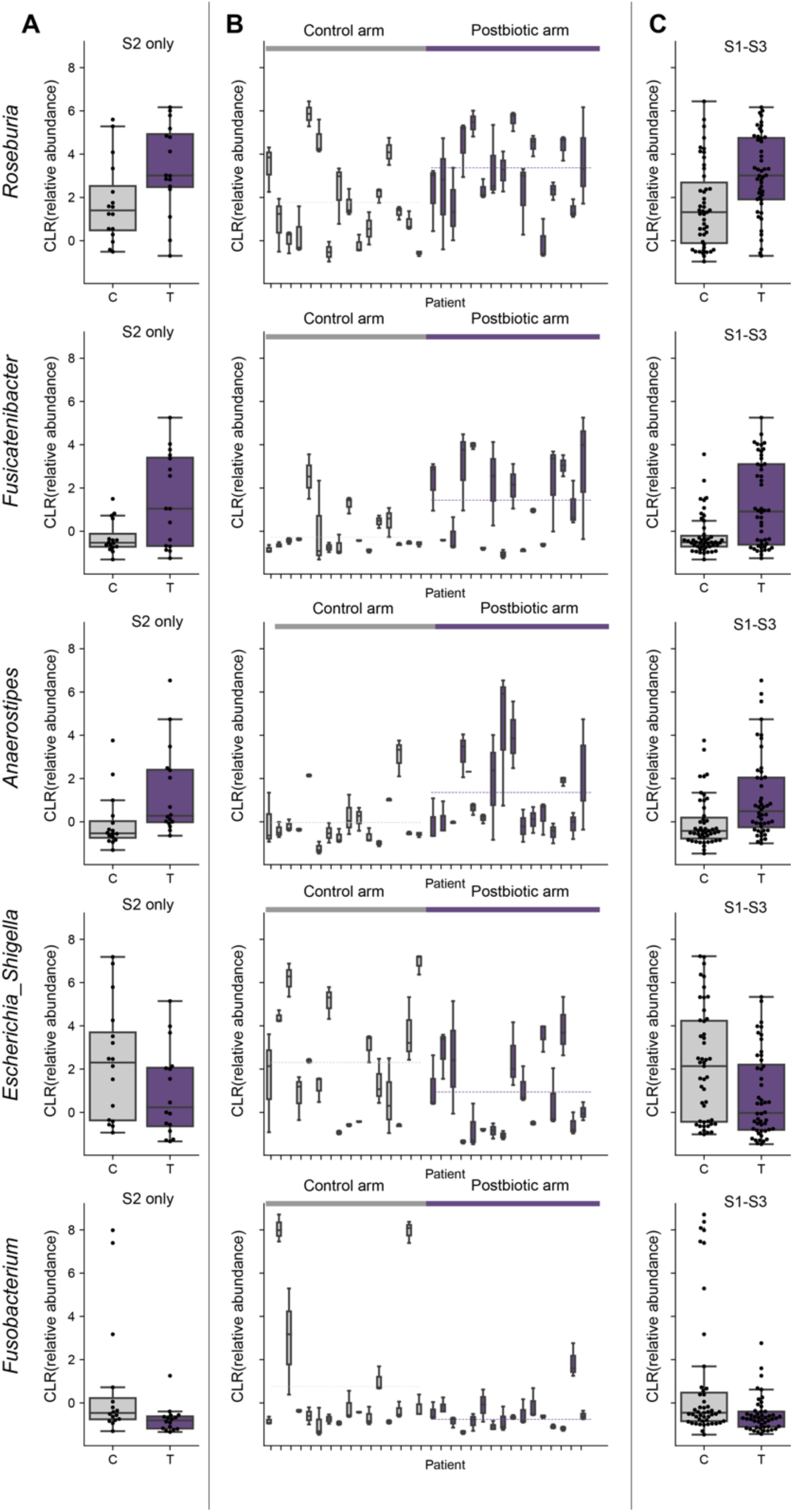
CLR transformed relative abundances of five most strongly associated genera across patients and time. Genera are organized in rows. **A**) CLR transformed relative abundances by treatment arm at timepoint S2 alone, **B)** for each patient in timepoints S1, S2, and S3 (dashed lines indicate medians per treatment arm), **C)** by treatment arm in timepoints S1, S2, and S3.

**Figure S5:**
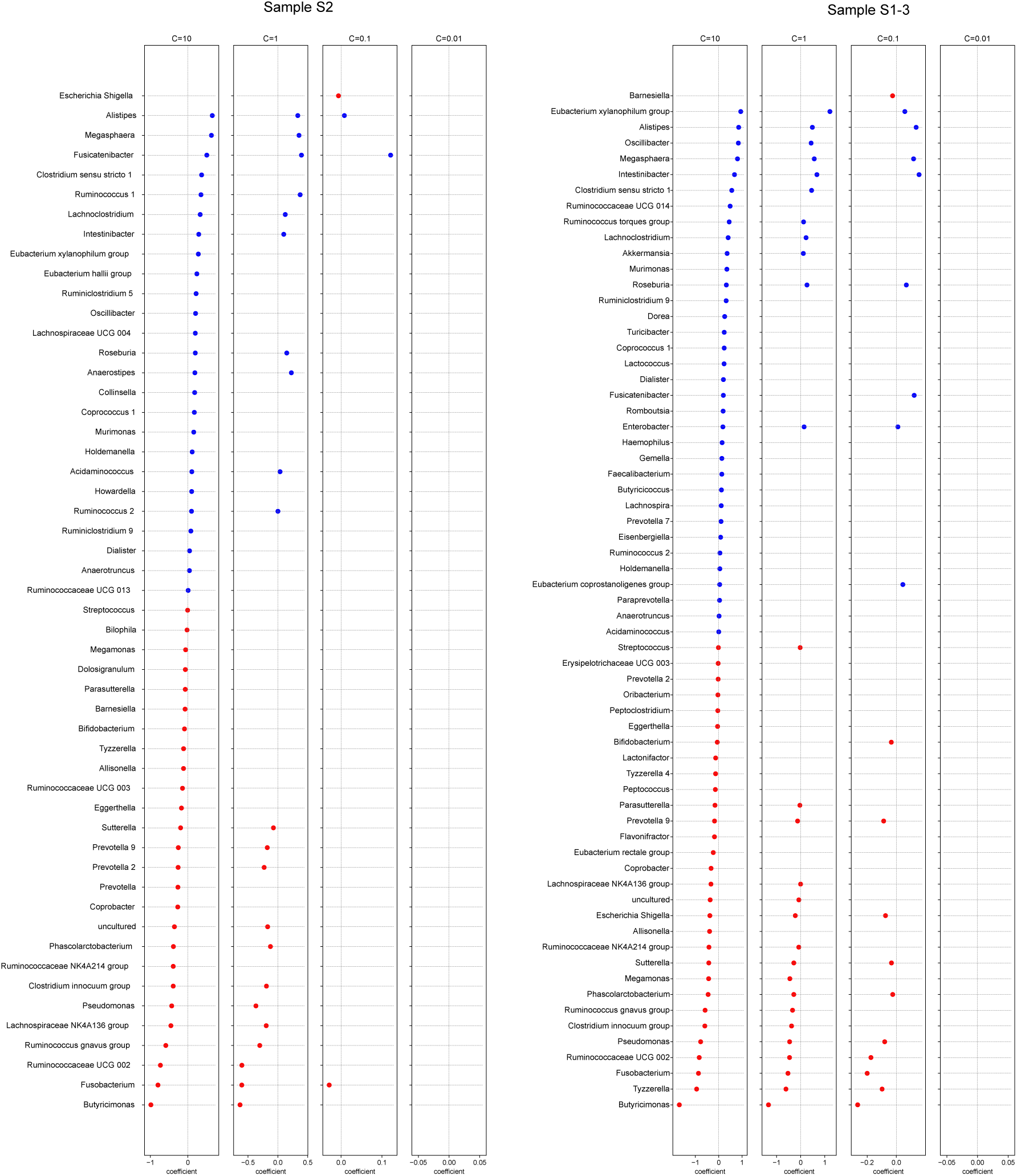
Multivariate logistic regression with varying regularization strengths (C) confirm univariate results. Coefficient estimates from L1-penalized logistic regressions on performed on CLR-transformed genus relative abundances in Sample S2 (**A**), or samples S1-S3 (**B**), at different regularization strengths up until all coefficients were set to zero (C: inverse regularization strength).

**Figure S6:**
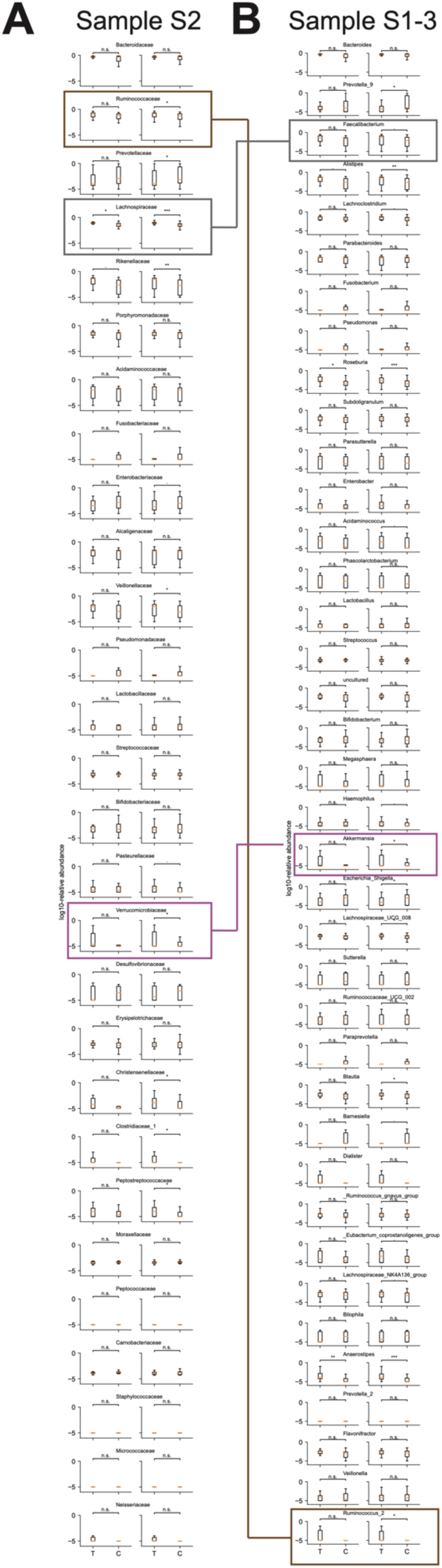
**Relative abundance of bacterial families and genera**, sorted by average abundance. Log-10 transformed relative family abundances in sample S2 (**A**) and genus (**B**) abundances pooled over samples S1-S3 in treated (T) and control (C) patients. Boxes highlight select taxa discussed as modulating cancer immunotherapies^31,32,35^. ***: p<0.001, **: p<0.01, *: p<0.05, .: p<0.1, n.s.: p>0.1; non-parametric Wilcoxon rank sums test not corrected for multiple hypotheses.

**Figure S7:**
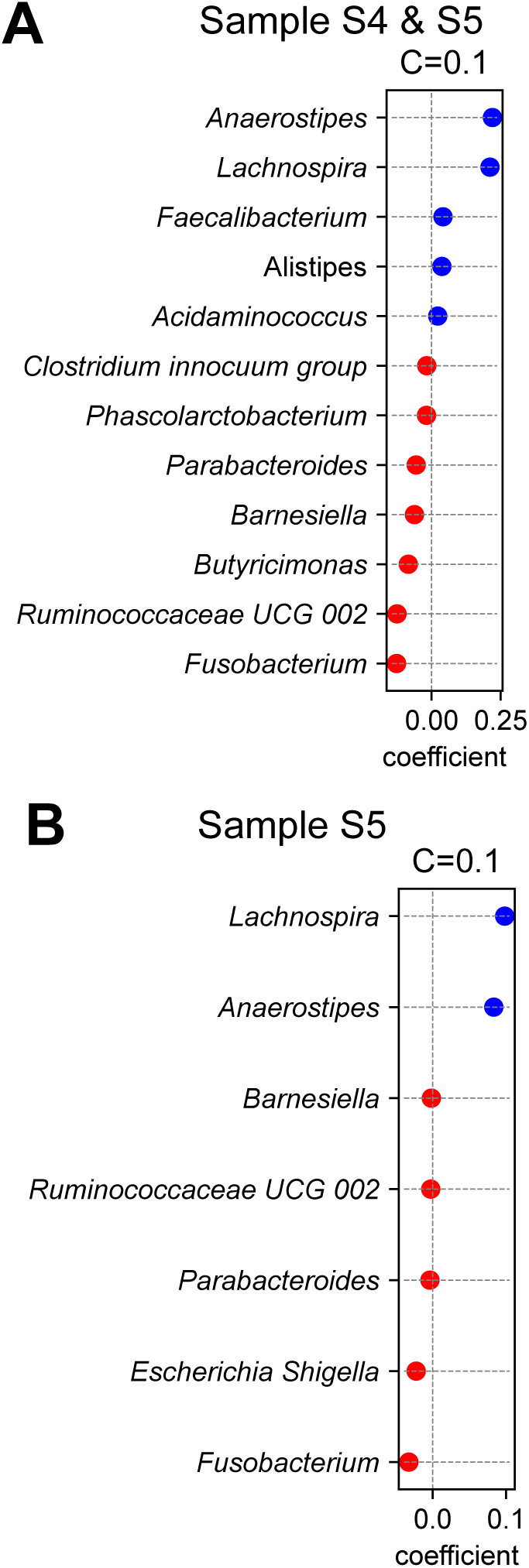
Multivariate L-1 regularized logistic regression of late samples. Coefficient estimates from L1-penalized logistic regressions on performed on CLR-transformed genus relative abundances in Sample S4 and S5 (**A**), or samples S5 alone (**B**) (C: inverse regularization strength).

**Figure S8:**
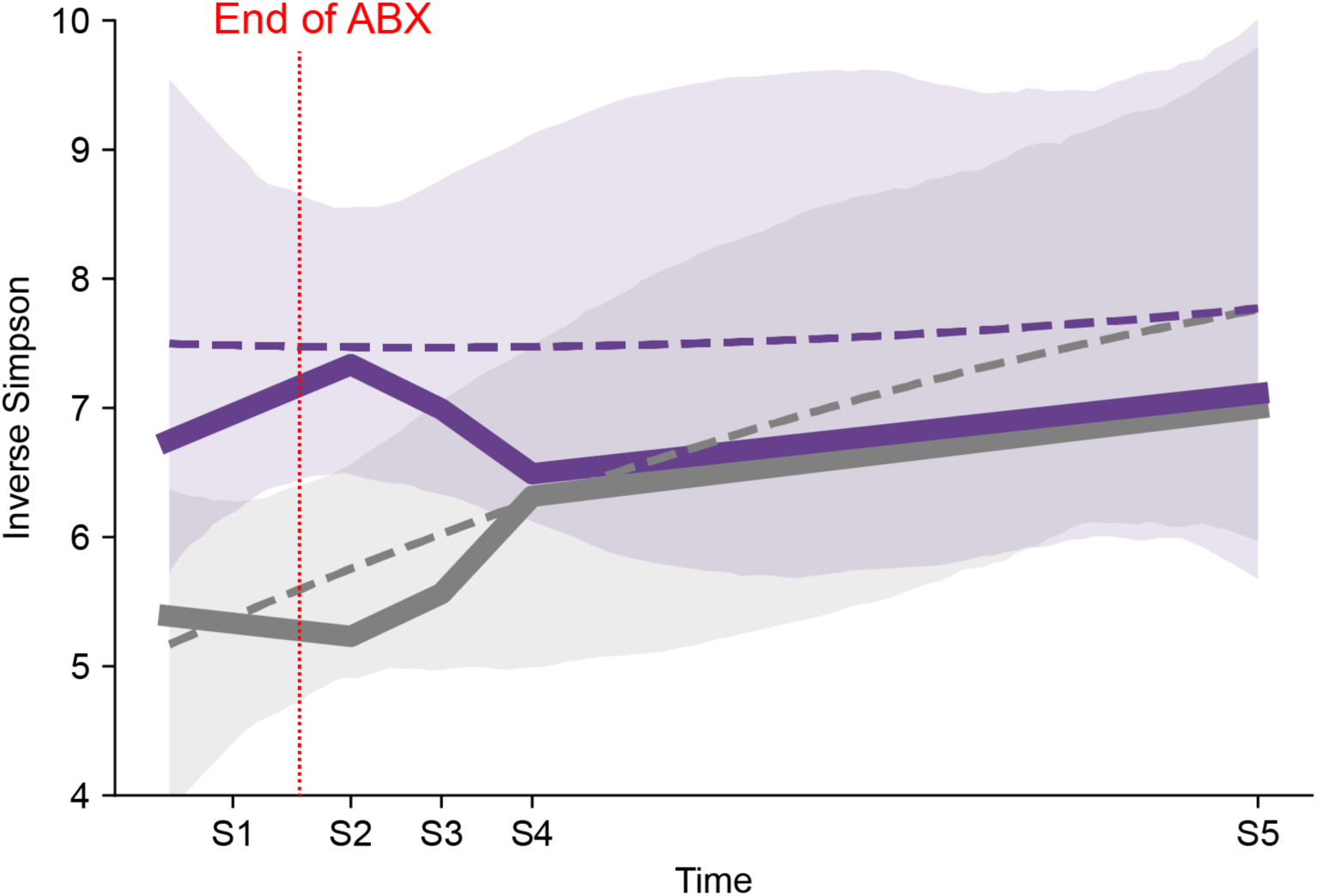
Bacterial alpha diversity is increased at the end of an antibiotic course in patients who received a postbiotic during their treatment and becomes similar ten days after finishing the antibiotic. Thick lines show a locally weighted scatterplot smoothing curve for treated (purple) and control (grey) patients’ bacterial alpha diversity across five longitudinally collected samples; dashed lines and shaded regions show mean and confidence intervals of a second-order line fit with time in days as predictor of alpha diversity.

**Table S1:**
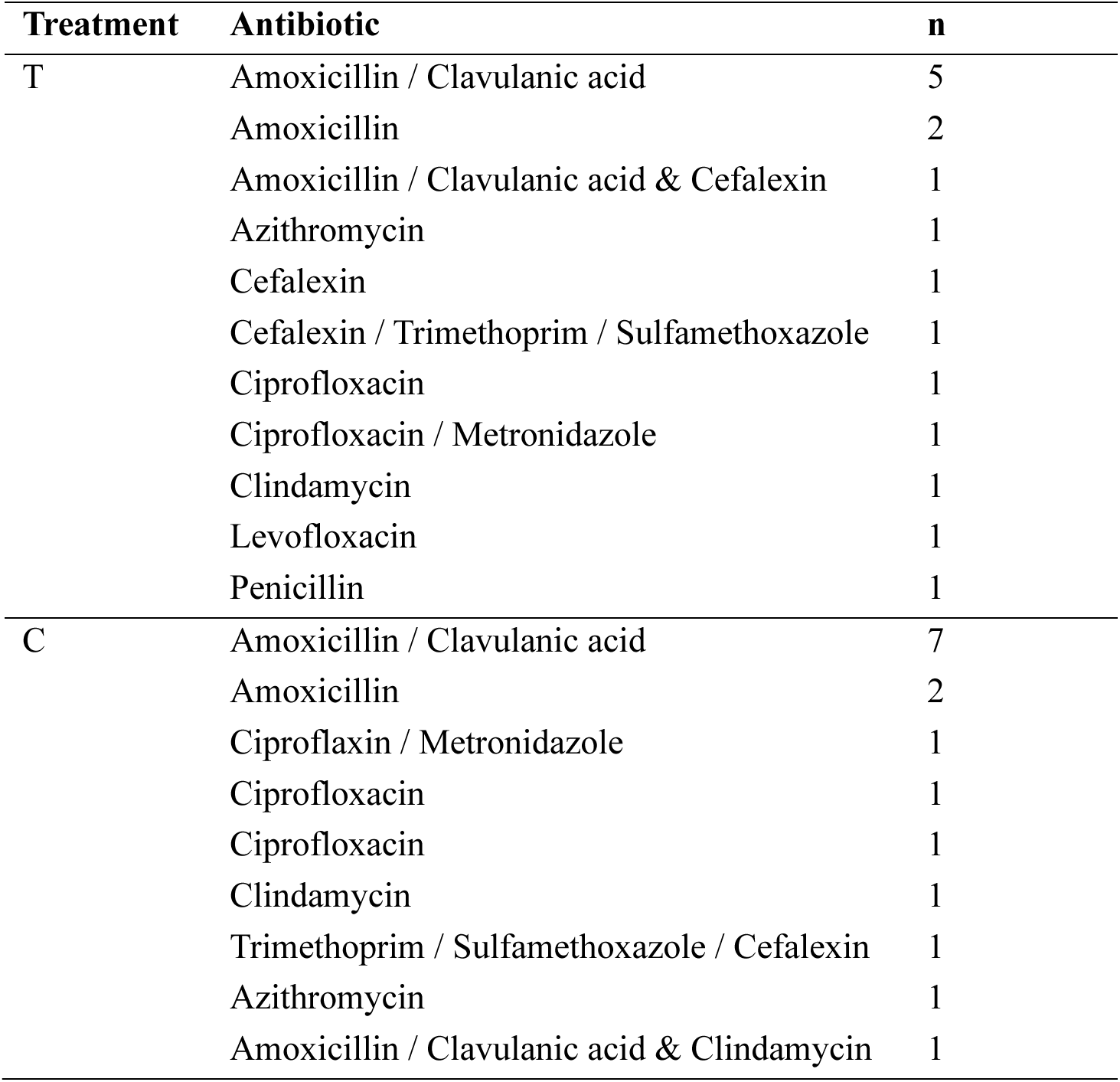
Antibiotic exposures by treatment arm.

**Table S2:**
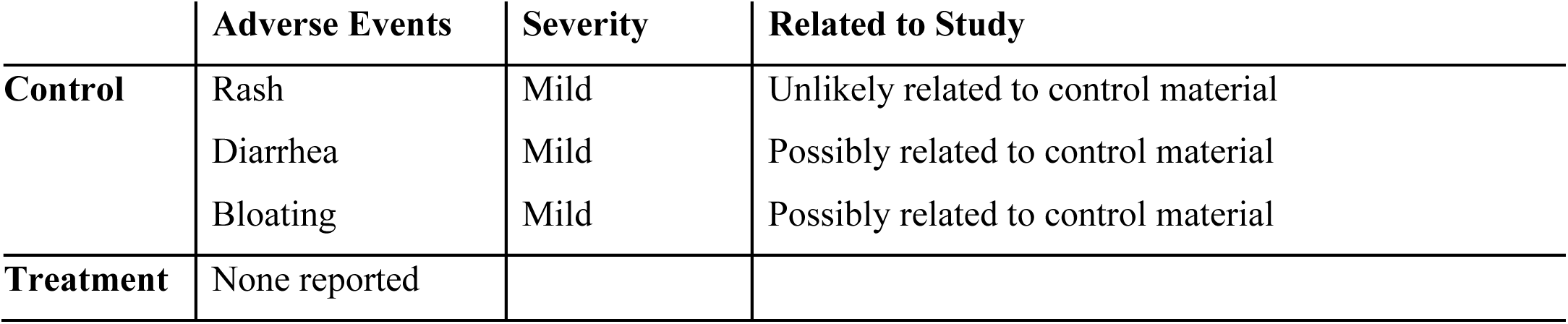
Reported adverse events.

**Table S3:** Data and statistical model output. Supplementary file “Table S3 Data and Statistics.xlsx”

## Notes

### Clinical Trial

Randomized controlled trial of PBP-GP-22 to affect microbiome composition; https://www.isrctn.com; trial registration ISRCTN30327931

### Author Declarations

This study protocol was reviewed and approved by the New England Institutional Review Board, and independent IRB located in Needham, MA (number: 120180088, trial registration ID: ISRCTN30327931.

