## Supplementary figures and images for "A randomized controlled trial of postbiotic administration during antibiotic treatment increases microbiome diversity and enriches health associated taxa"

### SuppFig1

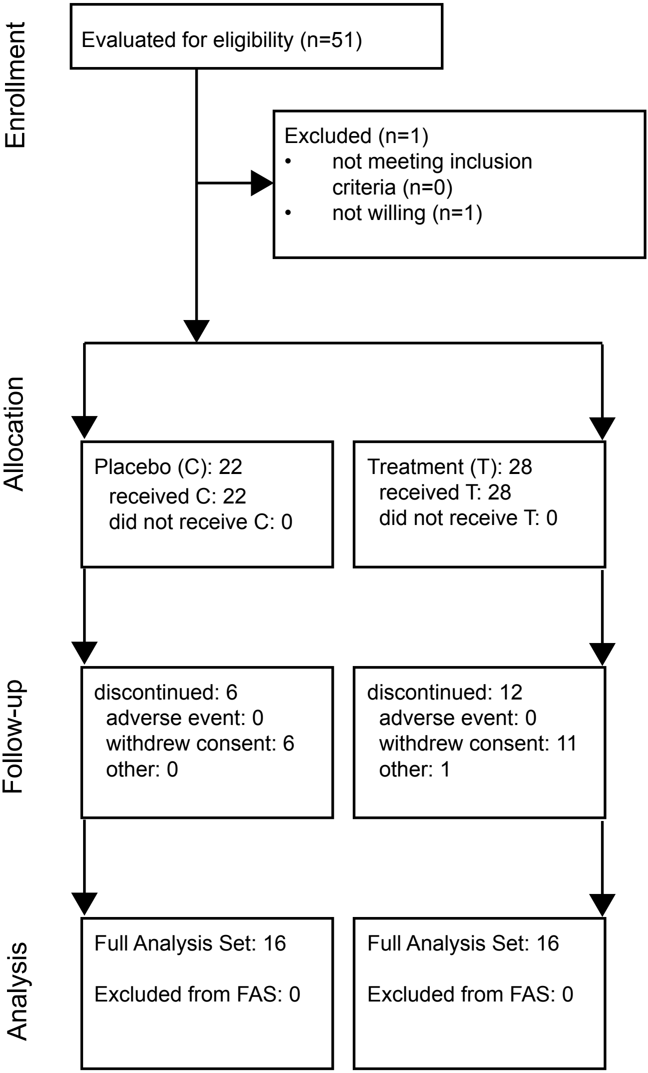

### SuppFig2

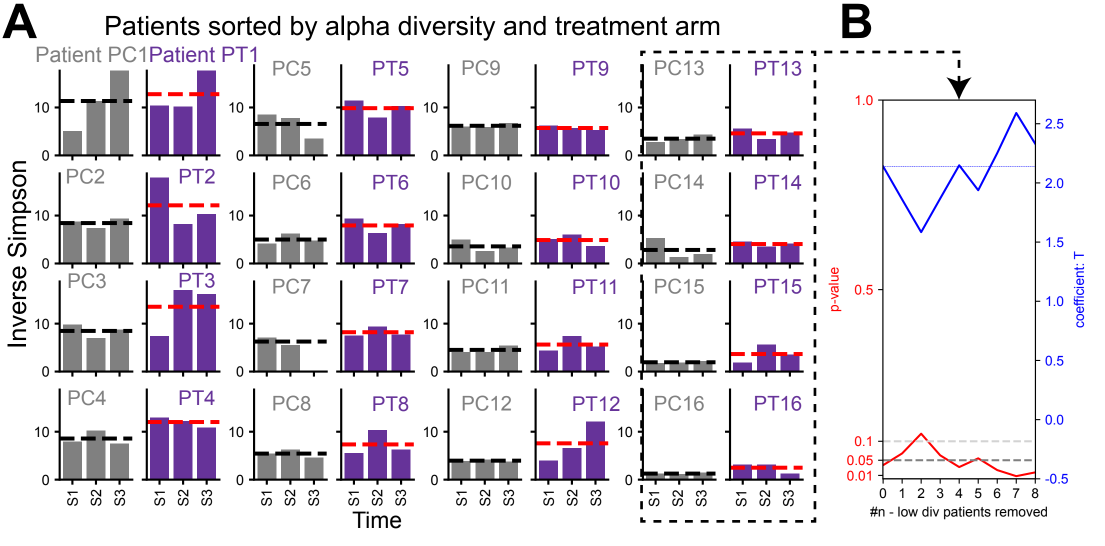

### SuppFig3

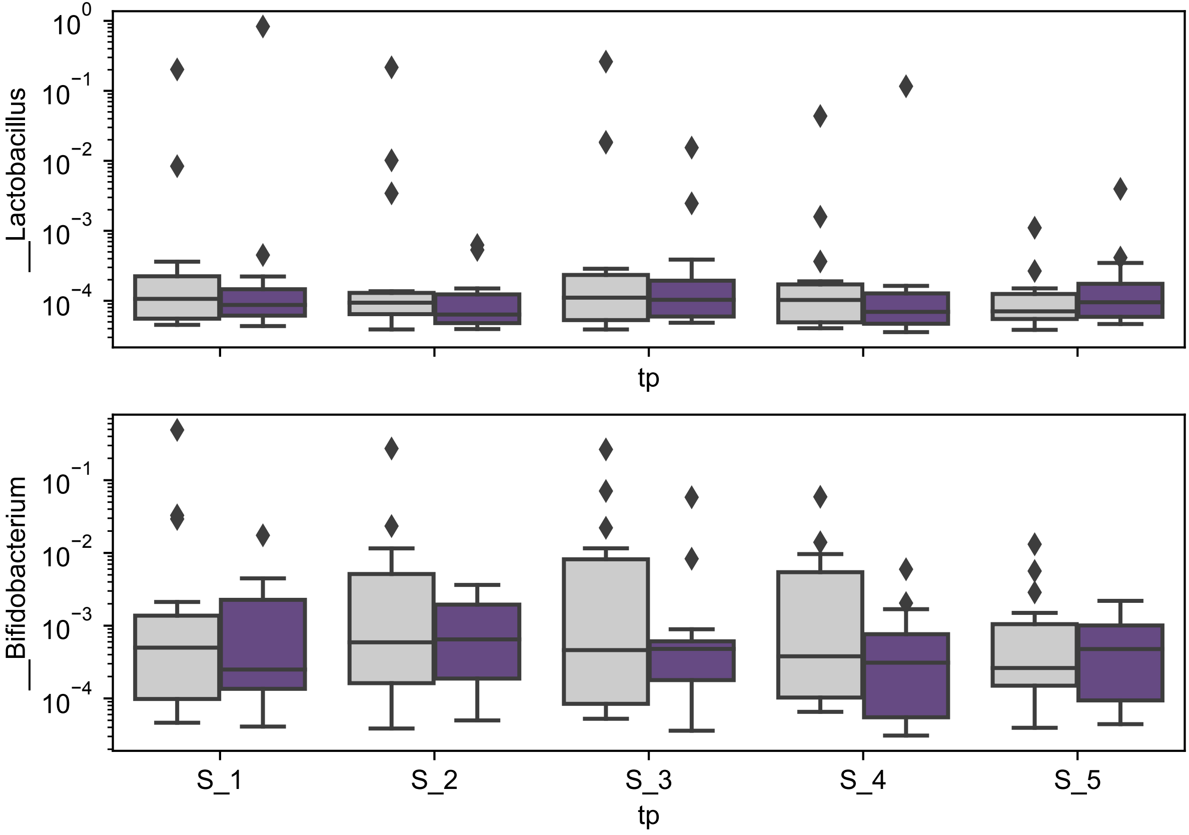

### SuppFig4

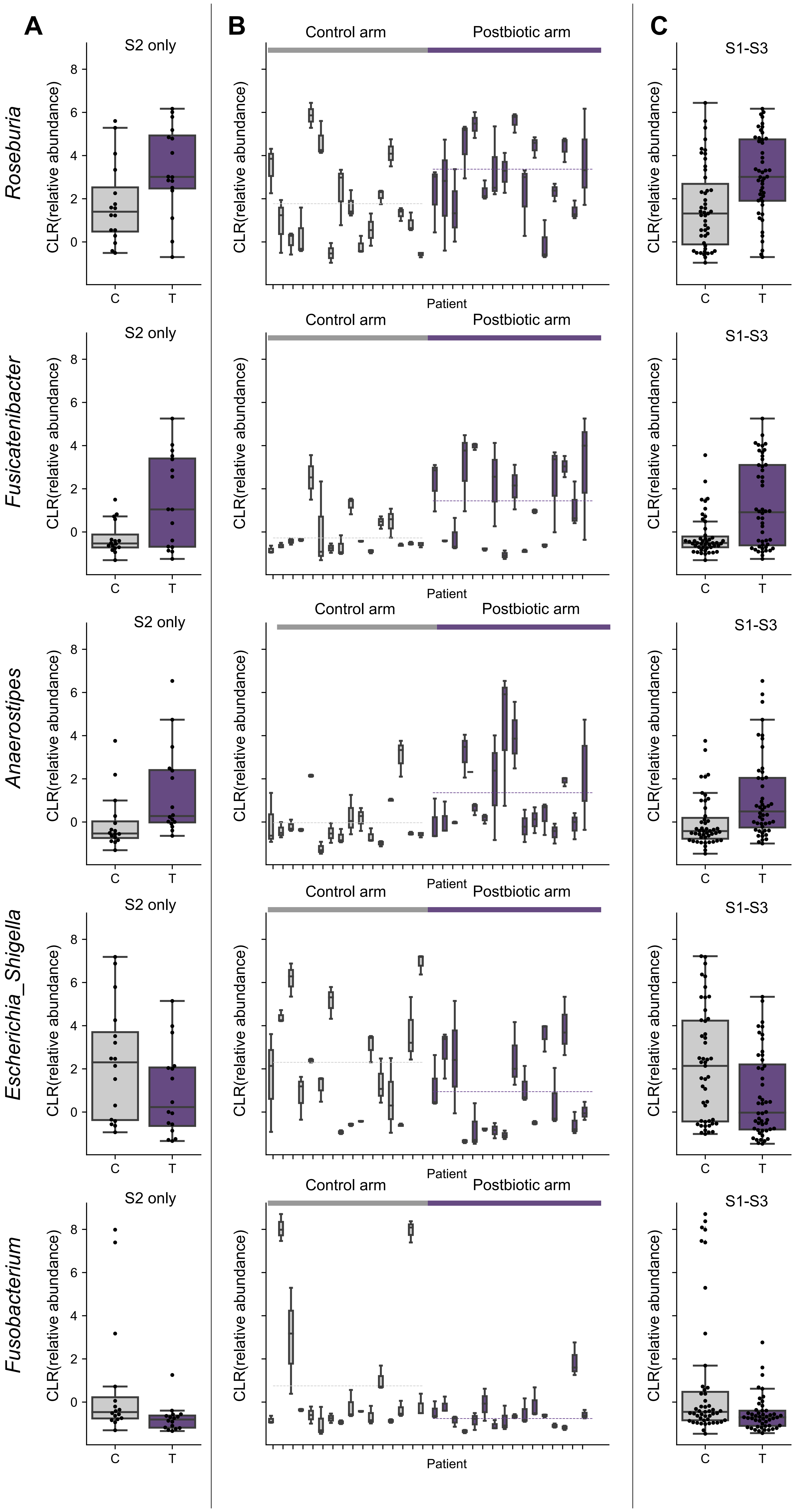

### SuppFig5

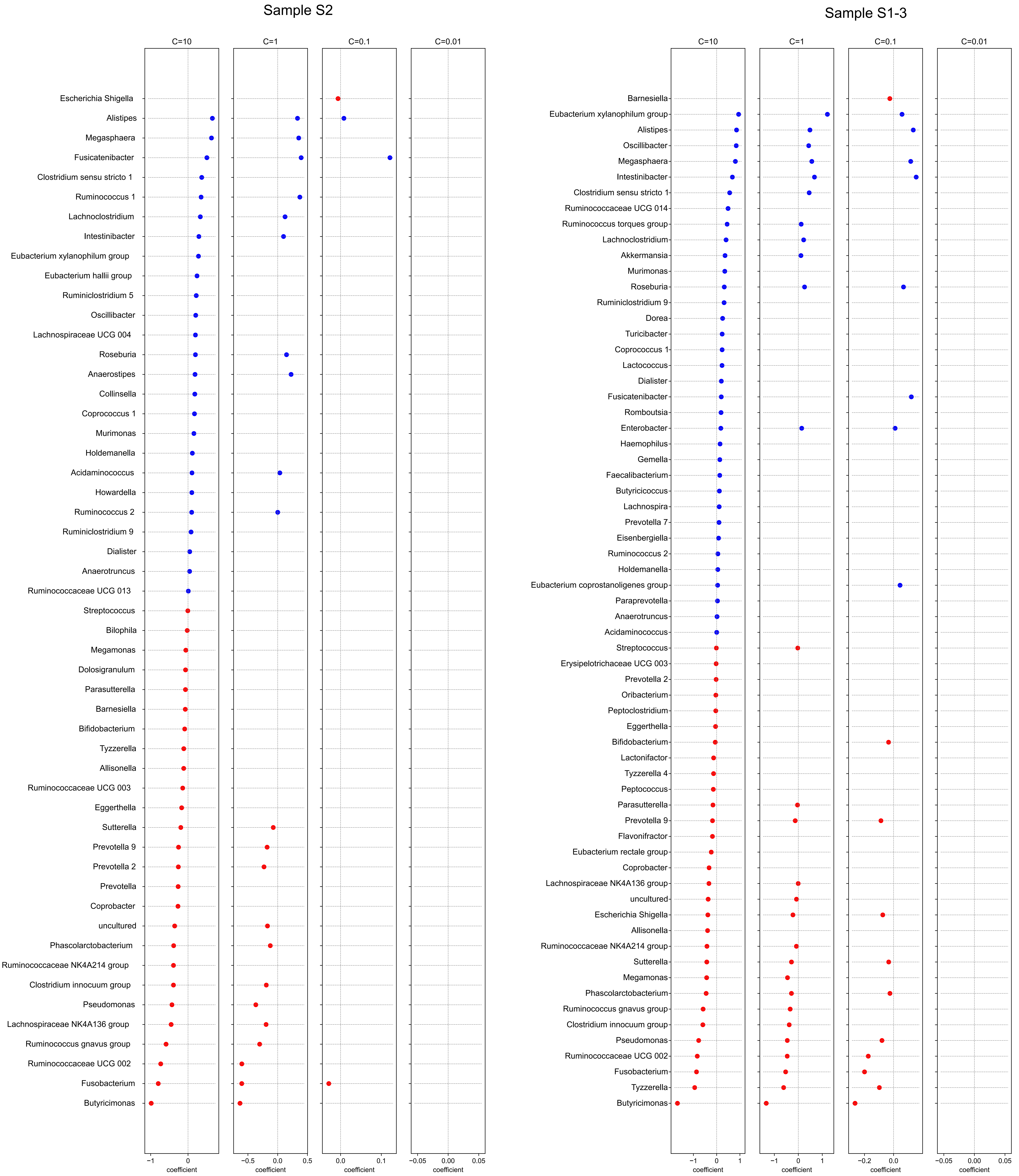

### SuppFig6

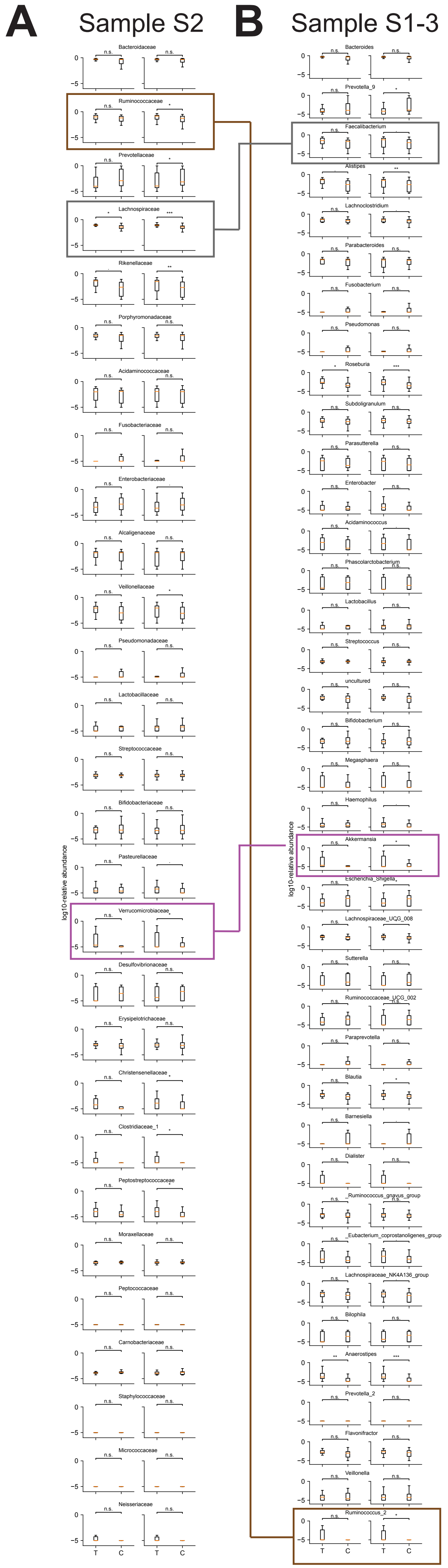

### SuppFig7

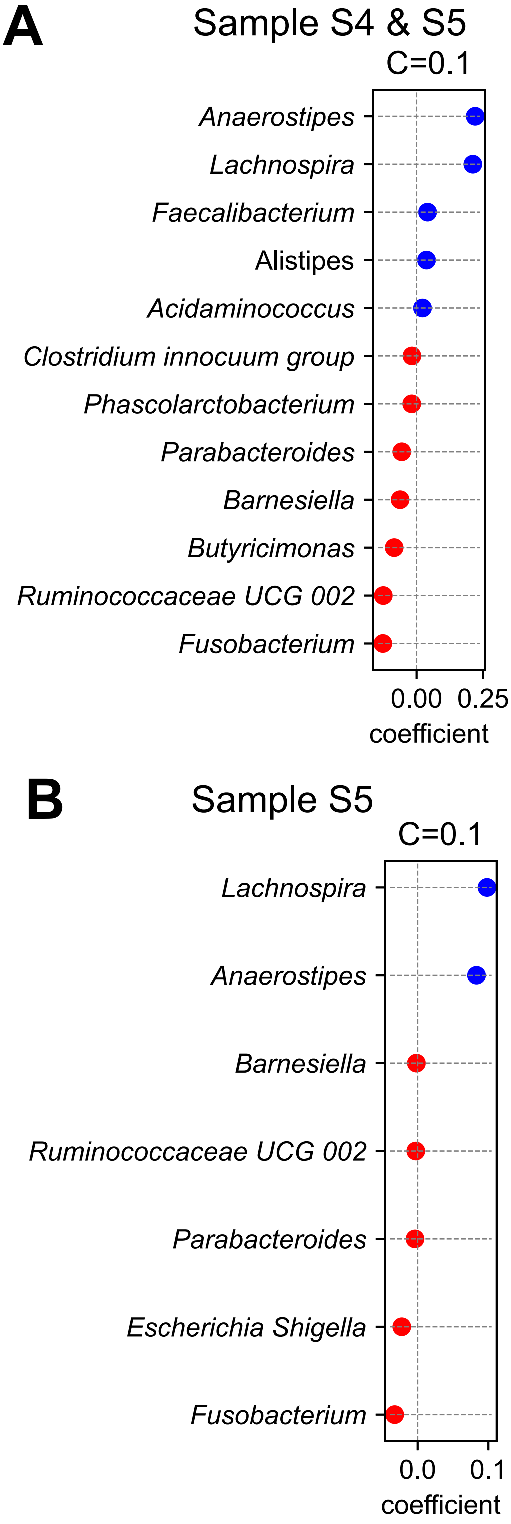

### SuppFig8

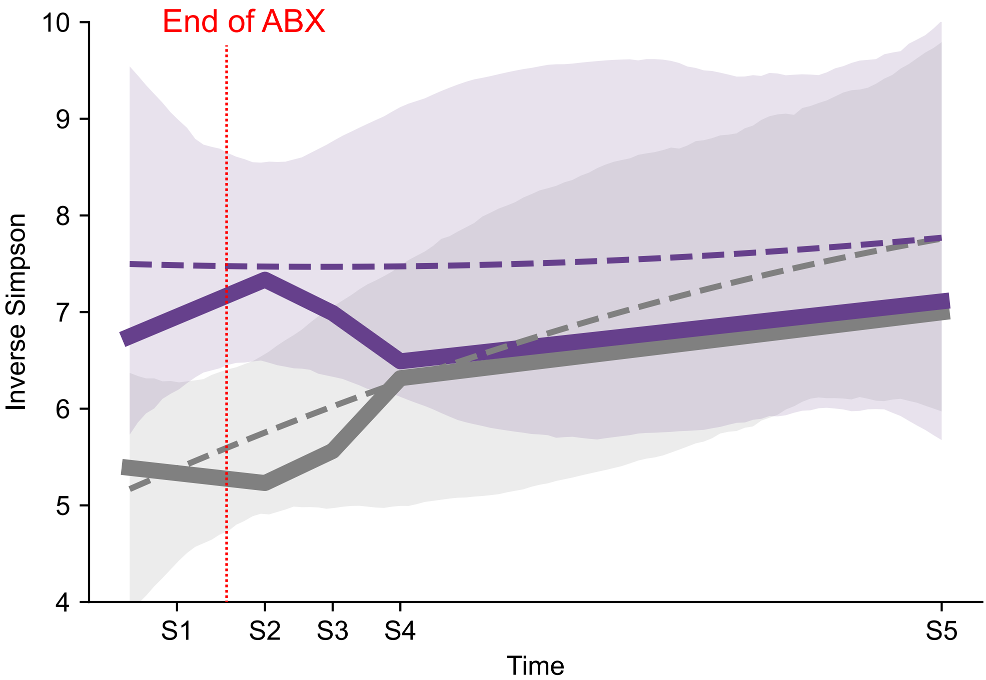
