## Supplementary material for "A randomized controlled trial of postbiotic administration during antibiotic treatment increases microbiome diversity and enriches health associated taxa": SuppMethods

Summary

================

The following is a summary of the pipeline used by Diversigen for processing sequencing runs targeting variable region four of the bacterial 16S rRNA gene. The pipeline is split into two modules: the first converts Illumina BCL files to fastq format, and the second clusters and classifies individual reads.

Process

================

Fastq Creation

----------------

The BCL files from a MiSeq run, in conjuction with a mapping file identifying samples associated with a particular molecular barcode are supplied to CASAVA, generating individual fastq files for each sample. A mismatch of one base pair is allowed, taking advantage of golay barcodes which require 3 mutations to collide with another barcode in use. These files are compressed using `pbzip2` for storage.

A second CASAVA run is initiated without the mapping file, producing three monolithic non-demultiplexed fastq for read 1 (forward), read 2 (barcode), and read 3 (reverse) for archival purposes. These files are compressed with `gzip` for storage.

Merging Paired Reads

----------------

The demultiplexed reads for a given collection of samples are taken together into this stage, to ensure clusters are consistent in the final output for that group of samples.

Fastq files containing the forward and reverse reads are merged using Usearch 7.0's `fastq_mergepairs` function, requiring to read pairs to overlap by at least 50 base pairs, have a merged length of at least 252 base pairs, a truncation quality above 5, and zero differences in the overlapping region.

These merged files are then filtered further, using Usearch 7.0's `fastq_filter` program. The final filter uses the strict fastq files, and only allows for a maximum expected error of 0.05. Merged reads at this point are relabeled to include the originating sample's name and combined into a single fastq file.

The fastq file is then filtered for PhiX using `bowtie2` and the `--very-sensitive` parameter setting, recording the percentage removed at this point. After removing PhiX reads, the fastq file is transformed into a fasta format file.

Read Clustering

----------------

1. The fasta file is run through Usearch 7.0's `derep_fulllength` program, creating a uc file, and a dereplicated fasta file.

2. The reads within the dereplicated fasta file are then sorted by size using Usearch 7.0's `sortbysize` program, and placed into a new fasta file.

3. At increments of 0.4%, the sequences are run through Usearch 7.0's `cluster_otus`, creating a clustered fasta file at each increment. These files are then filtered for any chimeras found by the program, and all chimera sequences are logged into their own file. The decisions made by the clustering program are also logged in a file.

4. The output from the previous increment is then fed into the next iteration, until a maximum of 3.2% is reached.

5. After the final run through the above loop, the final output is run through Usearch 7.0's `uchime_ref` program against the Gold database, using only the plus strand and allowing for no chimeras to create a file with no chimeras.

6. This new file is run through Usearch 7.0's `usearch_global` against the current Silva database, specifying ID to be set at 96.8%, using the plus strand, 0 maxaccepts and rejects.

7. The previously created dereplicated fasta file is then searched for any singletons, which are stripped from this file and placed into a separate singleton fasta file.

8. Using Usearch 7.0's `usearch_global` program, the singleton fasta file is then compared to the previously created sorted fasta file, using only the plus strand, allowing for 32 maxaccepts and 128 maxrejects and requiring an identity value of 99%. The results are stored.

9. All the files created in the loop so far are then run through a program developed in-house that resolves the iterative uparse steps, creating an OTU table, removing the chimera and singleton reads, and using the Silva database to map them.

10. The biom file is then summarized, and the statistics for the number of reads per sample that were mapped are recorded. This file with the statistics is then merged with a file that was generated for the overall read statistics, to give a file that shows the number of raw reads, and mapped versus unmapped reads per sample.

Data Exporting

----------------

After the loop itself has finished, all reads are concatenated into monolithic read files for read 1, read 2, and a combined reads fastq file. The final step in the process is to recover the barcodes reads for any raw data analysis. The monolithic fastqs are run through an in-house program that recovers these reads for delivery.

At this point, all files required are moved into the appropriate deliverables folder.

As a final step, a custom R script is called to summarize the biom file in an Excel workbook.

Results

================

The final files that are created are a monolithic read 1 and read 2 file, a barcodes file for both the raw and standard merge, a monolithic read file of the standard merge, an OTU table and associated stats sheet, and an example metadata mapping file.

List of Programs Used

================

* Bowtie2 (http://bowtie-bio.sourceforge.net/bowtie2/index.shtml)

* GNU Parallel (https://www.gnu.org/software/parallel/)

* pbzip2 (http://compression.ca/pbzip2/)

* Perl v5.16.2 (https://metacpan.org/release/RJBS/perl-5.16.2)

* R v3.0.2 (https://cran.r-project.org/bin/windows/base/old/3.0.2/)

* Silva v128 (https://www.arb-silva.de/documentation/release-128/)

* Usearch v7.0.1090 (http://www.drive5.com/Usearch/)

* Vegan v2.4-2 (https://github.com/vegandevs/vegan)
