## Supplementary material for "A randomized controlled trial of postbiotic administration during antibiotic treatment increases microbiome diversity and enriches health associated taxa": SuppTab1

**Table S1:** Antibiotic exposures by treatment arm

| Treatment | Antibiotic | n |
| --- | --- | --- |
| T | Amoxicillin / Clavulanic acid | 5 |
|  | Amoxicillin | 2 |
|  | Amoxicillin / Clavulanic acid & Cefalexin | 1 |
|  | Azithromycin | 1 |
|  | Cefalexin | 1 |
|  | Cefalexin / Trimethoprim / Sulfamethoxazole | 1 |
|  | Ciprofloxacin | 1 |
|  | Ciprofloxacin / Metronidazole | 1 |
|  | Clindamycin | 1 |
|  | Levofloxacin | 1 |
|  | Penicillin | 1 |
| C | Amoxicillin / Clavulanic acid | 7 |
|  | Amoxicillin | 2 |
|  | Ciproflaxin / Metronidazole | 1 |
|  | Ciprofloxacin | 1 |
|  | Ciprofloxacin | 1 |
|  | Clindamycin | 1 |
|  | Trimethoprim / Sulfamethoxazole / Cefalexin | 1 |
|  | Azithromycin | 1 |
|  | Amoxicillin / Clavulanic acid & Clindamycin | 1 |
