## Supplementary material for "A randomized controlled trial of postbiotic administration during antibiotic treatment increases microbiome diversity and enriches health associated taxa": SuppTab2

**Table S2: Reported adverse events**

|  | **Adverse Events** | **Severity** | **Related to Study** |
| --- | --- | --- | --- |
| **Control** | Rash | Mild | Unlikely related to control material |
|  | Diarrhea | Mild | Possibly related to control material |
|  | Bloating | Mild | Possibly related to control material |
| **Treatment** | None reported |  |  |
